# Descriptive Analysis of SARS-CoV-2 Genomics Data from Ambulatory Patients

**DOI:** 10.1101/2023.05.03.23289106

**Authors:** Nalini Ambrose, Alpesh Amin, Brian Anderson, Monica Bertagnolli, Francis Campion, Daniel Chow, Ashley Drews, Heath Farris, Fraser Gaspar, Stephen Jones, Tonia Korves, Bert Lopansri, James Musser, Eric Neumann, John O’Horo, Steven Piantadosi, Bobbi Pritt, Raymund Razonable, Seth Roberts, Suzanne Sandmeyer, David Stein, Farhaan Vahidy, Webb Brandon, Jennifer Yttri

## Abstract

**Background:** The COVID-19 pandemic has been characterized by ongoing evolution of the severe acute respiratory syndrome coronavirus 2 (SARS-CoV-2), with concomitant variation in viral transmissibility and morbidity. Within specific timeframes and geographic areas, multiple SARS-CoV-2 variants have coexisted in the human population, each characterized by distinct biologic and clinical features, such as varying susceptibility to neutralizing monoclonal antibodies (nMAbs), a major frontline treatment. As part of an observational real-world data study of the effectiveness of nMAbs for treatment of COVID-19, SARS-CoV-2 viral samples were obtained from patients under treatment, generating paired clinical and genomics data. This paper describes the processing pipeline and findings from the genomics portion of this combined data set.

**Methods:** SARS-CoV-2 sequences were generated from 14,796 diagnostic samples from four large U.S. health systems between July 2020 and March 2022. Among nMAbs-treated patients, samples were collected on the same day as, or prior to, treatment with nMAbs. Thus, these samples represent a snapshot of SARS-CoV-2 variants circulating in the respective patient groups, as opposed to variants that arose in response to specific treatments.

Health systems collected viral samples and performed library creation and sequencing according to local protocols, using tiled ARTIC amplicon primers. FASTQ files were submitted to a study data platform and processed through a common pipeline. This pipeline enabled a unified approach to quality control, assembly, and production of genomics features for downstream analysis.

**Results:** Alpha and pre-Alpha SARS-CoV-2 lineages were predominant in the data set prior to June 2021. From June 2021 through November 2021, Delta was the dominant variant. Beginning in December 2021, Omicron was dominant. A variety of mutations associated with decreased nMAbs binding to the spike protein *in vitro* were detected, including lineage-defining mutations and non-lineage-defining mutations such as E340A, G446V, and S494P. Distinct patterns of sequence gaps and ambiguous base calls were associated with distinct variants.

**Conclusions:** The distribution of SARS-CoV-2 variants, per WHO nomenclature, across epochs in this data set matched concurrent CDC genomic surveillance results across the U.S. Detection of putative nMAbs escape mutations within clinical samples was consistent with FDA decisions to amend EUAs as variants emerged. This genomics data set provides an opportunity to examine associations between SARS-CoV-2 genomic variation and clinical outcomes in the associated EHR data set. The expansion of real-world data sets such as this to study the relationship between viral sequence and treatment outcomes could provide the foundation for future efforts to achieve near-real-time understanding of clinical outcomes related to genomic variation over time, and evidence to update treatment decisions more rapidly and to greater effect during ongoing and future pandemics.

## INTRODUCTION

Since the severe acute respiratory syndrome coronavirus 2 (SARS-CoV-2) first emerged within the human population in late 2019, efforts have been made worldwide to surveil the emergence of novel genetic lineages of the virus. The course of the pandemic has been marked by significant evolution of SARS-CoV-2, with different variants characterized by distinct transmissibility,^1^ morbidity,^2^ and prevalence across time.^3^

As part of a large observational real-world evidence (RWE) study of the effectiveness of neutralizing monoclonal antibodies (nMAbs) directed toward SARS-CoV-2, patient-level clinical data and linked SARS-CoV-2 genomics sequence data from diagnostic samples were collected from four geographically diverse health systems within the United States.^4^ Analyses of this clinical data to date have found that the nMAbs treatments in use during the study period (bamlanivimab, bamlanivimab-etesevimab, casirivimab-imdevimab, and sotrovimab) were effective in reducing hospitalization and mortality. When genomic variation at the level of WHO variant was incorporated, nMAbs treatments were found to be effective in patients infected with Delta, but not in those infected with Omicron, variants.^4^

This paper presents genomics methods and results from 14,796 SARS-CoV-2 viral samples obtained as part of the RWE nMAbs study, including the distribution of WHO variants over time, nucleotide diversity, and putative nMAbs escape mutations within the samples. The unusual aspects of this data set include genomics data that were 1) from samples collected across a broad time range (July 2020 through March 2022); 2) from samples collected at four geographically distinct health systems within the United States; and 3) analyzed via a common pipeline with stringent quality controls, beginning with FASTQ files as inputs and producing a variety of outputs, including genomics features such as viral lineage label and mutation frequencies.

## METHODS

Clinical and viral genomics data were submitted to a secure, centralized registry by four participating health systems as part of an observational study of the effectiveness of nMAbs. A detailed description of the patient population, including inclusion/exclusion criteria for the RWE nMAbs study and patient clinical and other characteristics can be found in the companion article.^4^ Patients were eligible for inclusion in this data set if they had a laboratory-confirmed diagnosis of COVID-19 and at least one FDA emergency use authorization (EUA) criterion for high risk of poor outcome. Clinical data were submitted for 196,571 patients; after applying the inclusion and exclusion criteria, the final clinical cohort included data from 167,183 patients, of whom 15.1% (25,241) were treated with nMAbs.

The same four health systems submitted SARS-CoV-2 viral genomics data derived from patient biologic samples, obtained between July 2020 and March 2022. Health systems identified available samples collected during clinical practice for sequencing. Increased proportions of samples from the Delta and Omicron timeframes were requested for inclusion in the study data set, to provide additional insights into these variants. Additionally, health systems submitted genomic sequences most likely to meet established data quality criteria for inclusion. Health systems were not equally represented in the data set due to differences in sequencing capacity and availability of patient samples. Data from a total of 16,055 samples were submitted to the centralized registry. After filtering for data quality and other issues, viral genomics data from 14,796 samples remained, with one sample per patient, all taken on the day of COVID-19 diagnosis; results presented in this report refer to this set unless otherwise noted. A subset of 13,703 of these samples belonged to patients who were included in the final RWE nMAbs clinical cohort and thus had definitive determination of nMAb treatment status. Details of pipeline and filtering for data quality and other issues follow.

Health systems, or in one case an associated public health department, performed sample storage, library creation, and sequencing according to local protocols. Among treated patients, most samples (93%) were definitively taken prior to treatment; in the remaining cases, diagnostic samples were taken the same day as treatment. All sites used tiled, multiplexed primers published by the ARTIC network (versions 3 and 4).^5^ Sequencing was performed with Illumina machines. A total of 24,887 FASTQ files (7,223 files from single-end read runs and 17,664 paired files from paired-end read runs) representing 16,055 viral samples from 16,033 distinct patients were shared to the centralized registry.

Within the centralized registry, a parallelized pipeline was created to produce genomics features and drive downstream analytics while imposing uniform quality constraints. All FASTQ files were scrubbed of human reads, including human-specific controls from the ARTIC protocol, using the National Center for Biotechnology Information Sequence Read Archive human read removal tool.^6^ Regular expressions applied across the FASTQ metadata identified potential protected health information and personally identifiable information for removal. In subsequent pipeline steps, paired FASTQ files from paired-end sequencing runs were processed together and FASTQ files from single-end sequencing runs were processed individually.

FASTQ files were quality trimmed and adapters removed using fastp,^7^ specifying a threshold Phred score of 30, an unqualified percentage limit of 20, and a minimum read length of 15. The reads from pre-processed FASTQ files were then aligned to the reference SARS-CoV-2 genome (NCBI Reference Sequence NC_045512.2) using minimap2,^8^ resulting in a BAM file for each sample. The PCR primers were soft-clipped from these BAM files using iVar trim,^9^ specifying a minimum quality threshold of 30 and a minimum read length to retain after trimming of 30, and including reads with no primers.

The primer-trimmed BAM files were next processed by SAMTOOLS to create pileup files for each sample using SAMTOOLS mpileup.^10^ The pileup files were inputs to iVar variants^9^ for variant calling, specifying a minimum quality score of 30, 0.75 as the minimum frequency threshold to call variants, and a minimum read depth of 10. SnpEff^11^ was used to annotate the called variants, including classification of nucleotide variants as high effect (e.g., frame shift deletion, stop gain), moderate (e.g., in-frame deletion, missense mutation), or low (e.g., synonymous mutation). Additionally, the pileup files were used to make consensus FASTA files for each sample using iVar consensus^9^ with a minimum quality score of 30, 0.75 as the minimum frequency threshold to call variants, and a minimum read depth of 10. The consensus FASTAs were used as inputs to the pangolin tool^12^ (PANGO designations 1.3 and 1.6, both from April 2022)^13^ to produce viral lineage calls for each sample. PANGO lineages were mapped to equivalent WHO variants^14^ to enable coarser categorization and facilitate interpretation.

To identify the position of gaps and ambiguous base calls in each sample relative to the reference genome, consensus FASTA files were aligned to the reference genome using mummer4^15^ (nucmer and show-aligns scripts). The resulting simple text-based alignment file was parsed and the relevant positions stored for each sample. Custom scripts (R version 4.2.1 and Python version 3.6) were developed to assess the presence or absence of putative nMAb strong escape mutations using data and criteria from the J. Bloom laboratory^16^ and based on the pipeline variant calls and the positions of ambiguous base calls and gaps. The nMAbs investigated were bamlanivimab, etesevimab, casirivimab, imdevimab, and sotrovimab, which were in clinical use for treatment during the time of the study, and cilgavimab and tixagevimab, which were in prophylactic use.

Data representing each sample were subjected to multiple data quality filters based on findings from multiple pipeline steps. Data from samples were retained for analysis if they met the following criteria: 1) consensus FASTA sequence length >= 27,000 base pairs, 2) number of ambiguous base calls^17,18^ <= 5,000, 3) rate of base calls with a quality Phred score of at least 30 >= 80%, and 4) the percentage of the genome covered at least 10X >= 90% (or >= 80% if criteria 1, 2, and 3 were all satisfied). Additional checks to provide further context were performed, including proportion of reads aligned, average guanine-cytosine (GC) content, RT-PCR cycle threshold (CT), and variant call coverage. RT-PCR CT was inconsistently reported by health systems (missing for 71% of samples) and therefore was not used as an exclusionary data quality criterion. For patients with replicate samples passing data quality thresholds, one sample was selected for inclusion from among samples collected on the day of COVID-19 diagnosis, based on lower RT-PRC cycle threshold (CT) value (if reported) or arbitrarily (if CT not reported).

Data from the RWE nMAbs study, including deidentified clinical data, FASTQ files with viral genomic sequences, and a table of viral genomics features associated with each patient (e.g., PANGO lineage, presence/absence of putative escape mutations to specific nMAbs) were submitted to the National COVID-19 Cohort Collaborative.

## RESULTS

Genomics data from 16,055 samples, taken between July 2020 and March 2022, were passed through the data processing pipeline. A total of 1,042 samples failed one or more of the critical data quality checks, most commonly the check on percentage of the genome covered at a minimum of 10X depth (n = 1,032). Of the 15,013 samples passing critical data quality checks, 94 could not be linked to patient clinical data and were removed from analyses. A small number of the remaining samples were not taken on the day of initial COVID-19 diagnosis (n = 117) or represented replicate samples from the same patient (n = 6). This paper details the results and findings for the remaining 14,796 samples that passed all data quality checks, could be linked to patient clinical data, were taken on the day of initial COVID-19 diagnosis, and were not replicate samples. A subset of 13,703 samples were from patients who met all clinical inclusion and exclusion criteria for the broader RWE study of nMAb effectiveness;^4^ wherever treatment status is discussed below (i.e., treated with nMAbs vs. not treated), the total number of samples under consideration is 13,703.

Patients with viral genomics data form a subset of the larger cohort used in the RWE study of nMAb safety and effectiveness. Table 1 is a comparison of demographic and clinical characteristics of the group of patients with and without associated viral genomics data. The proportion of patients with viral genomics data who were treated with nMAb is larger than in the group of patients without viral genomics data (30.2% and 12.6%, respectively). Aside from this, the profile of demographic features and comorbidities for these two groups is similar.

**Table 1.** Comparison of Clinical Characteristics of Patients with and without Viral Genomic Samples. The total number of patients considered (196,571) represents all patients for whom clinical data was submitted to the RWE nMAbs central registry.

| Characteristic | Genomic Sample<br>Not Available,<br>n = 181,775 | Genomic Sample<br>Available,<br>n = 14,796 |
| --- | --- | --- |
| Treated with nMAb | 22,868 (12.6%) | 4,467 (30.2%) |
| Age in years – mean (standard deviation) | 47.4 (18.6) | 48.4 (18.3) |
| Gender female | 103,732 (57.1%) | 8,330 (56.3%) |
| Gender not specified | 7 (<0.1%) | 0 (0.0%) |
| Race American Indian or Alaska Native | 1,545 (0.8%) | 121 (0.8%) |
| Race Asian | 5,266 (2.9%) | 393 (2.7%) |
| Race Black or African American | 13,507 (7.4%) | 885 (6.0%) |
| Race Native Hawaiian or Other Pacific Islander | 1,871 (1.0%) | 127 (0.9%) |
| Race White | 148,148 (81.5%) | 12,256 (82.8%) |
| Race Other | 7,294 (4.0%) | 771 (5.2%) |
| Race Not specified | 4,144 (2.3%) | 243 (1.6%) |
| Ethnicity Hispanic or Latino | 29,264 (16.1%) | 2,195 (14.8%) |
| Ethnicity not specified | 5,143 (2.8%) | 384 (2.6%) |
| Smoker | 11,791 (6.5%) | 987 (6.7%) |
| Pregnant | 5,556 (3.1%) | 434 (2.9%) |
| Comorbidities/Coexisting Conditions |  |  |
| Obesity | 76,484 (42.1%) | 6,736 (45.5%) |
| Hypertension | 38,989 (21.4%) | 3,269 (22.1%) |
| Diabetes mellitus | 20,037 (11.0%) | 1,694 (11.4%) |
| Chronic pulmonary disease | 16,958 (9.3%) | 1,392 (9.4%) |
| Thrombotic or hematological disorders | 13,055 (7.2%) | 1,118 (7.6%) |
| Hypothyroidism | 12,209 (6.7%) | 1,070 (7.2%) |
| Malignancy | 8,555 (4.7%) | 748 (5.1%) |
| Peripheral vascular disease | 8,264 (4.5%) | 721 (4.9%) |

Approved for Public Release; Distribution Unlimited. Public Release Case Number 22-2693 © 2023 The MITRE Corporation. All Rights Reserved.
| Characteristic | Genomic Sample<br>Not Available,<br>n = 181,775 | Genomic Sample<br>Available,<br>n = 14,796 |
| --- | --- | --- |
| Chronic kidney disease | 8,425 (4.6%) | 708 (4.8%) |
| Neurological disorders or paralysis | 7,629 (4.2%) | 603 (4.1%) |
| Chronic liver disease | 6,435 (3.5%) | 506 (3.4%) |
| Heart failure | 5,738 (3.2%) | 467 (3.2%) |
| Valvular heart disease | 3,710 (2.0%) | 267 (1.8%) |

Genomics samples were classified by WHO variant^14^ and PANGO lineage.^13^ Of the 14,796 genomic samples included in this analysis, 9,058 (61.2%) samples were identified as WHO variant Delta; 3,188 (21.5%) as Omicron; 1,107 (7.5%) as Alpha; 351 (2.4%) as Epsilon; 82 (0.55%) as Gamma; 68 (0.46%) as Mu; and less than 25 each as Iota, Beta, Zeta, Lambda, Eta, and Kappa. A total of 898 (6.1%) genomes were classified as belonging to PANGO lineages that do not belong to any of the WHO variants, meaning they descended from lineages that were common prior to the WHO nomenclature system. Overall, 196 different PANGO lineages, including 81 sub-lineages within Delta and 33 sub-lineages within Omicron, were identified (Table S1). Among the Omicron samples, 3,175 were identified as sub-variant BA.1 and 12 as BA.2. Table S2 shows counts of different PANGO lineages across time.

Most viral genomics samples were taken between July 2021 and January 2022 (Figure 1a). Samples taken from patients diagnosed prior to June 2021 represented a mix of Alpha, Epsilon, and “No WHO Equivalent” (i.e., lineages that were common prior to the WHO nomenclature system). Beginning in June 2021 and continuing through November 2021, the Delta variant was dominant (Figure 1b). From December 2021 forward, Omicron was the most prevalent variant.

**Figure 1a.**
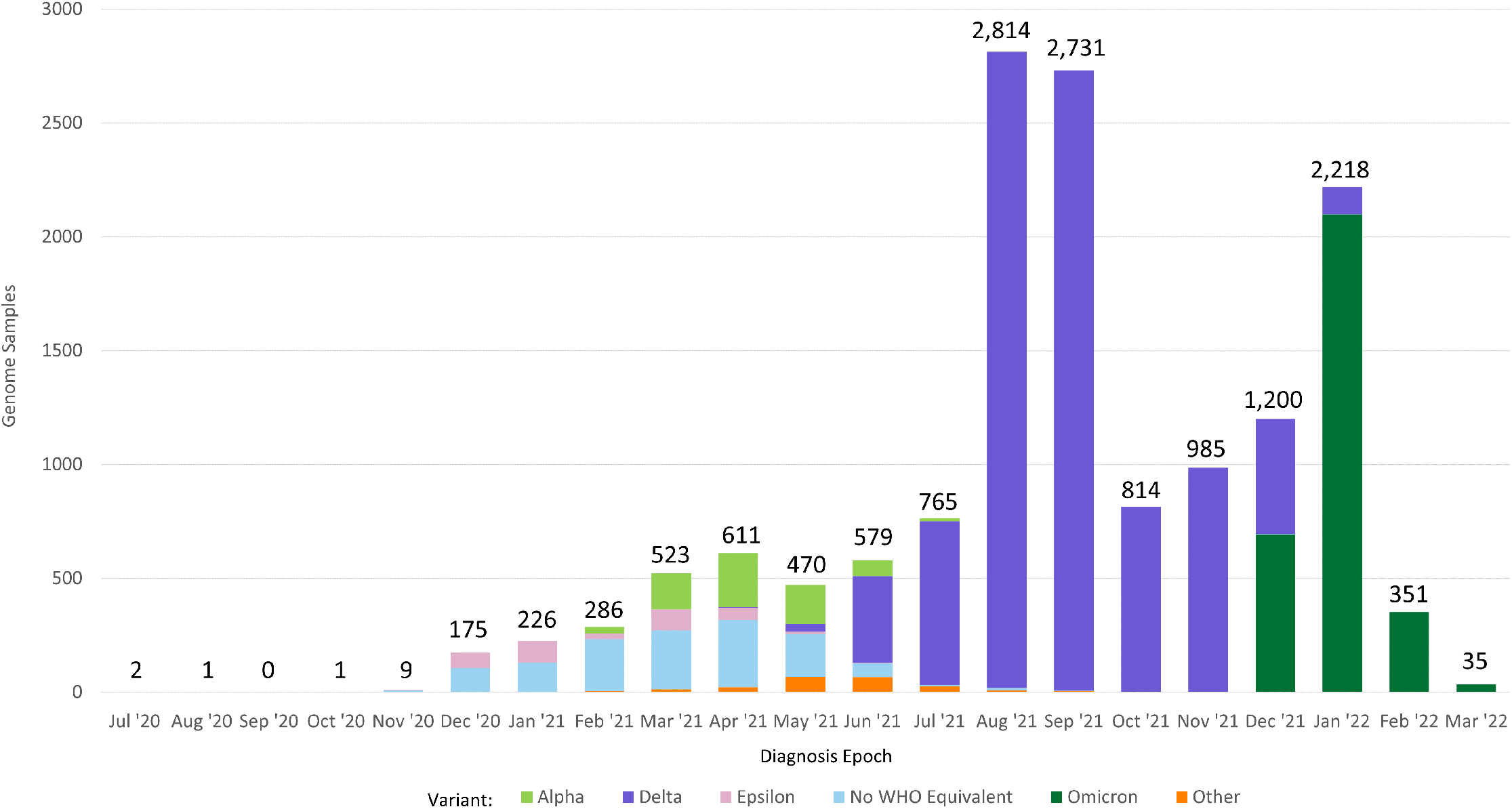
Distribution of Sequenced Genomic Samples by Diagnosis Epoch and WHO Variant.

**Figure 1b.**
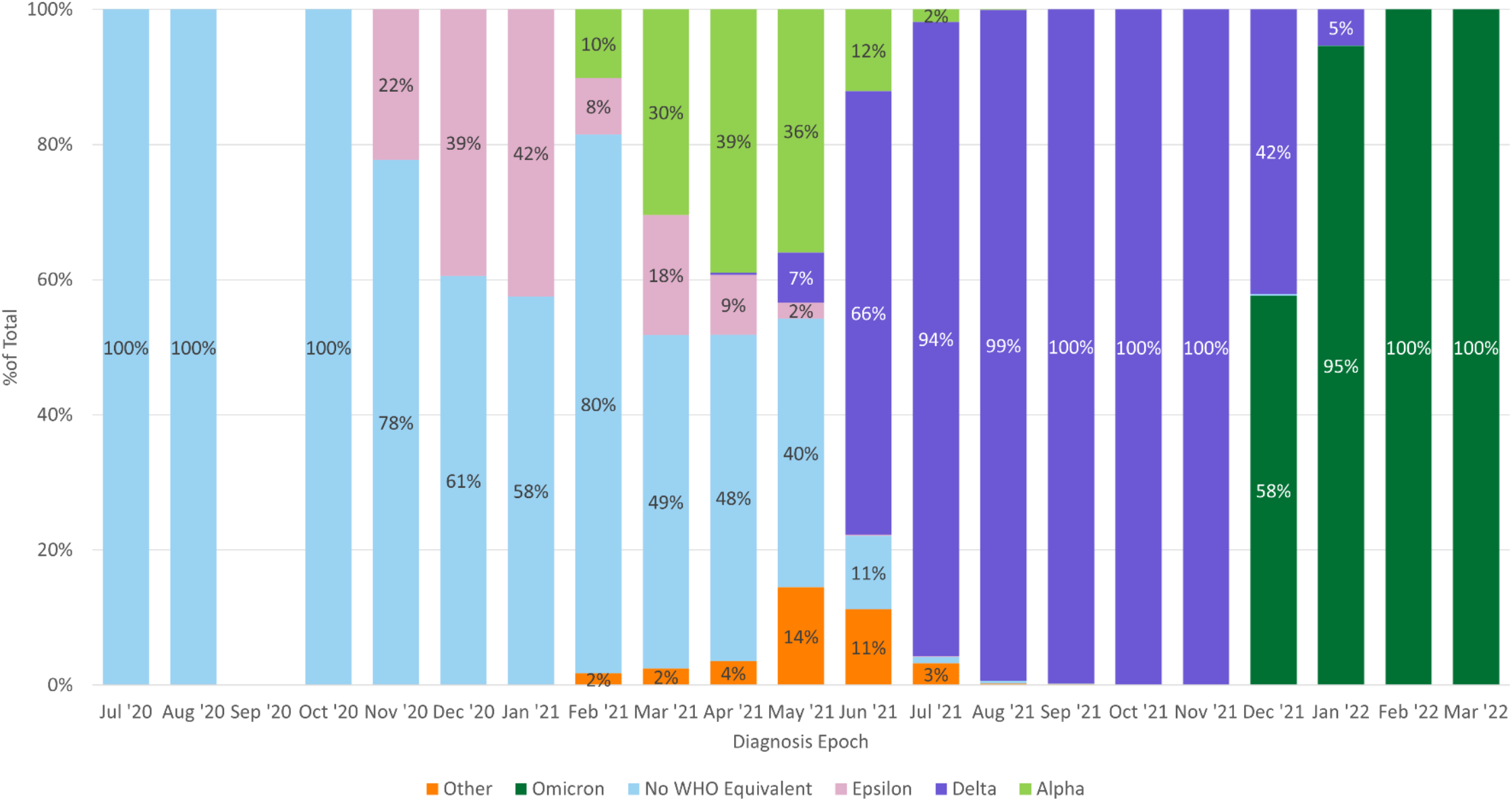
Percentage of WHO Variants in Sequenced Genomic Samples by Diagnosis Epoch. Stacked bars represent the number (1a) or proportion (1b) of sequences corresponding to WHO variants for each diagnosis epoch. “Other” categorizes variants represented in low numbers within the registry. Genome sequences were obtained for a subset of patients based on sample and sequencing reagent availability. There was a shortage of reagents in late 2021 and early 2022 that may have contributed to lower than expected numbers of Omicron sequences. The total number of samples represented is 14,796.

Across all genomic samples in the data set, 13,522 unique mutations were identified, including 12,861 (95%) SNPs and 661 (5%) insertions and deletions (indels). Mutations included 569 (4%) nucleotide variants predicted to have high effects on the virus (e.g., mutations causing premature protein truncation or frameshifts); 7,618 (56%) predicted to have moderate effects (SNPs causing amino acid changes and in-frame indels); 4,862 (36%) predicted to have low effects (SNPs within genes that do not alter amino acids); and 473 (3.5%) called as modifier effects (mutations outside of identified gene open reading frames).^11^ There were 133 unique nucleotide variants predicted to cause amino acid substitutions, insertions, or deletions in the spike protein (S protein) receptor-binding domain (RBD) region of sequences evaluated for escape variants. Figure 2 shows the distributions of mutations in the S protein within Delta and Omicron samples.

**Figure 2.**
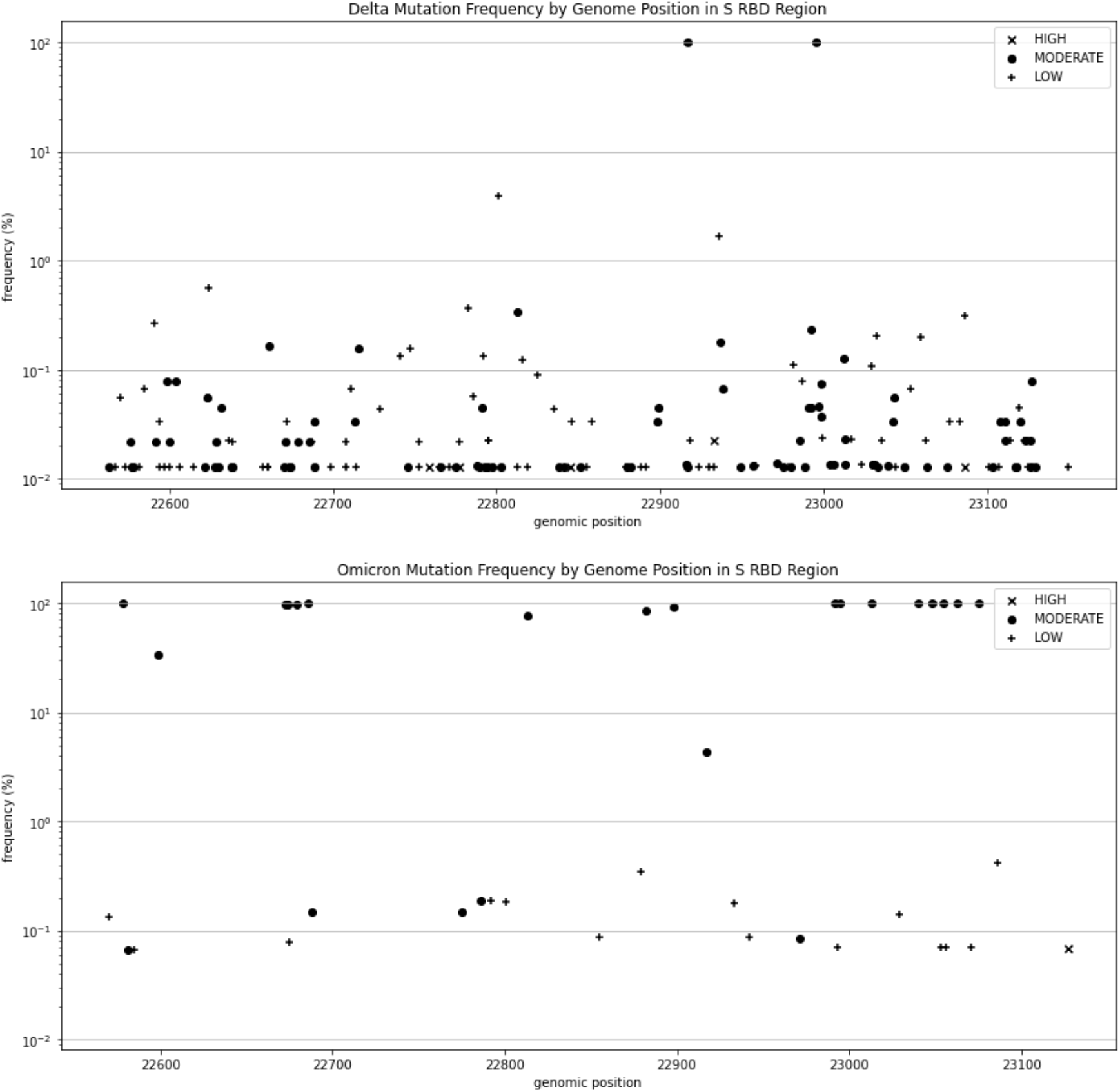
Nucleotide Variants in the S Protein RBD Region by Genome Position and Frequency within Delta (top) and Omicron (bottom) Samples. Predicted effects of nucleotide changes (low, moderate, high) are distinguished by the different markers, as shown in the legend. Frequencies were determined using only samples with unambiguous base calls in the corresponding locations.

Multiple putative nMAbs escape mutations were detected across the samples. Table 2 shows the detection of 24 unique amino acid substitutions in the S protein RBD region that in prior deep mutational scanning experiments decreased binding to the various nMAbs.^16,19^ While some of these mutations (e.g., D405N, K417N, L452R) were lineage defining, 13 were not.^14,19–23^ Twelve of the latter were found in five or fewer samples, and included P337S (in one Omicron sample, January 2022), E340A (in one Omicron sample, January 2022), R346S (in two Delta samples, December 2021), N440Y (in one Delta sample, January 2022), G446V (in one Epsilon and four Delta samples, February, July, August, November, and December 2021), Y473C (in one Delta sample, December 2021), A475V (in one Alpha sample, April 2021), G476D (in one Alpha sample, June 2021), E484V (in two Delta samples, August and October 2021), F490Y (in one Delta sample, November 2021), Q493E (in one Delta sample, October 2021), and P499R (in one Mu sample, September 2021). The mutation S494P was found in 32 samples across multiple WHO variants (No WHO Equivalent, Alpha, Epsilon, Delta, and Omicron) and epochs (in January through May, August, and December 2021 and February 2022).

**Table 2.**
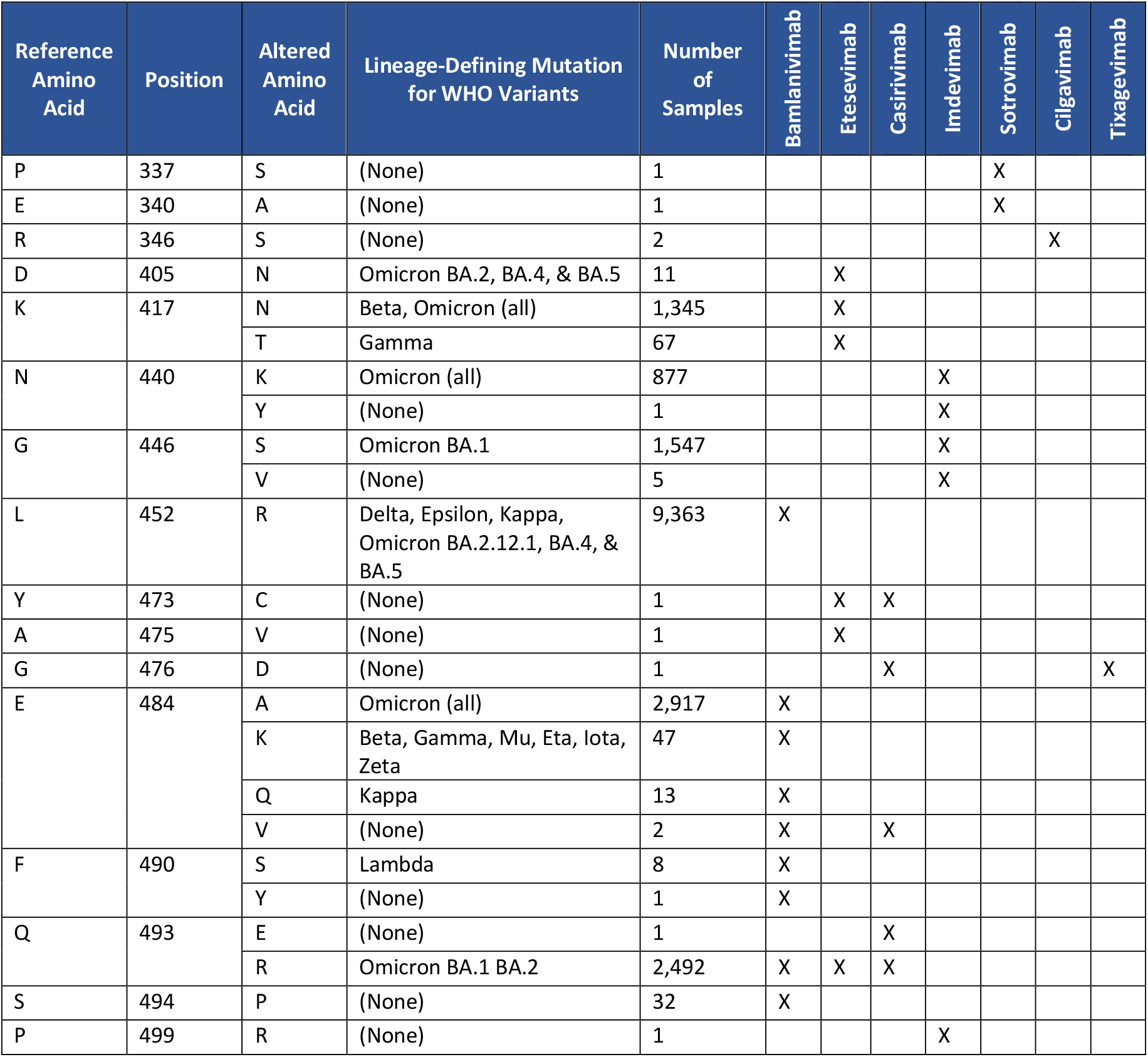
Detection of Putative mAb Escape Mutations in the Data Set. Presence of putative escape mutations is determined for the subset of samples with unambiguous base calls in the corresponding location.

In the non-Omicron samples, the occurrence of more than one putative escape mutation for an nMAb within a sample was rare. Among the 6,799 non-Omicron samples with complete sequence data in the S RBD region, 17 samples had two of the putative escape mutations for bamlanivimab (L452R and either E484A, E484Q, E484V, or S494P); none of the non-Omicron samples had more than one for the other nMAbs. Omicron has multiple characteristic mutations that are predicted to cause escape, including two putative escape mutations for each bamlanivimab, etesivimab, and imdevimab; Omicron BA.2 also has additional characteristic putative escape mutations for each bamlanivimab and etesivimab. While missing data in the S RBD region in many Omicron samples limited our ability to assess the presence of multiple putative escape per sample, among the 490 Omicron samples with complete sequence data in the RBD region, only one had a single putative escape mutation (S494P) beyond the lineage-defining ones.

In addition to the putative escape mutations from the deep mutational scanning data, analyses identified mutation S371L, which is characteristic of Omicron BA.1, and S371F, which is characteristic of Omicron BA.2 and later sub-variants; these mutations have been found to confer escape to multiple nMAbs despite not strongly reducing nMAb binding in deep mutational scanning experiments.^19^ Additional mutations highlighted by the CDC^24^ with potential but unknown impact on nMAbs were also seen within the data set: G339D was found within 3,060 sequences; S477N within 2938 sequences; T478K within 11,908 sequences; G496S within 2,919 sequences; Q498R within 2,954 sequences; N501Y within 4,276 sequences; and Y505H within 2,986 sequences.

There were relatively few instances where patients were treated with an nMAb while harboring virus with a putative escape mutation(s) to that particular nMAb (Figure 3). Among nMAb-treated patients, 176 had at least one putative escape mutation for each nMAb in their treatment; this represents ∼5% of the samples with no missing escape mutation data for the corresponding nMAb from treated patients. We explored the possibility of an analysis of clinical outcomes comparing patients with and without putative escape mutations for the nMAbs with which they were treated. There were insufficient instances to support such an analysis in depth. For example, Figure 3 reflects that one of the best opportunities for such an analysis would have been patients treated with bamlanivimab-etesevimab in December 2021; of the 124 patients treated with bamlanivimab-etesevimab and having viral genomics data in this month, 76 were infected by a SARS-CoV-2 virus with putative escape mutations for both bamlanivimab and etesevimab, while the remaining 48 were not. We observed only two patients with a 14-day hospitalization in the former group and none in the latter.

**Figure 3.**
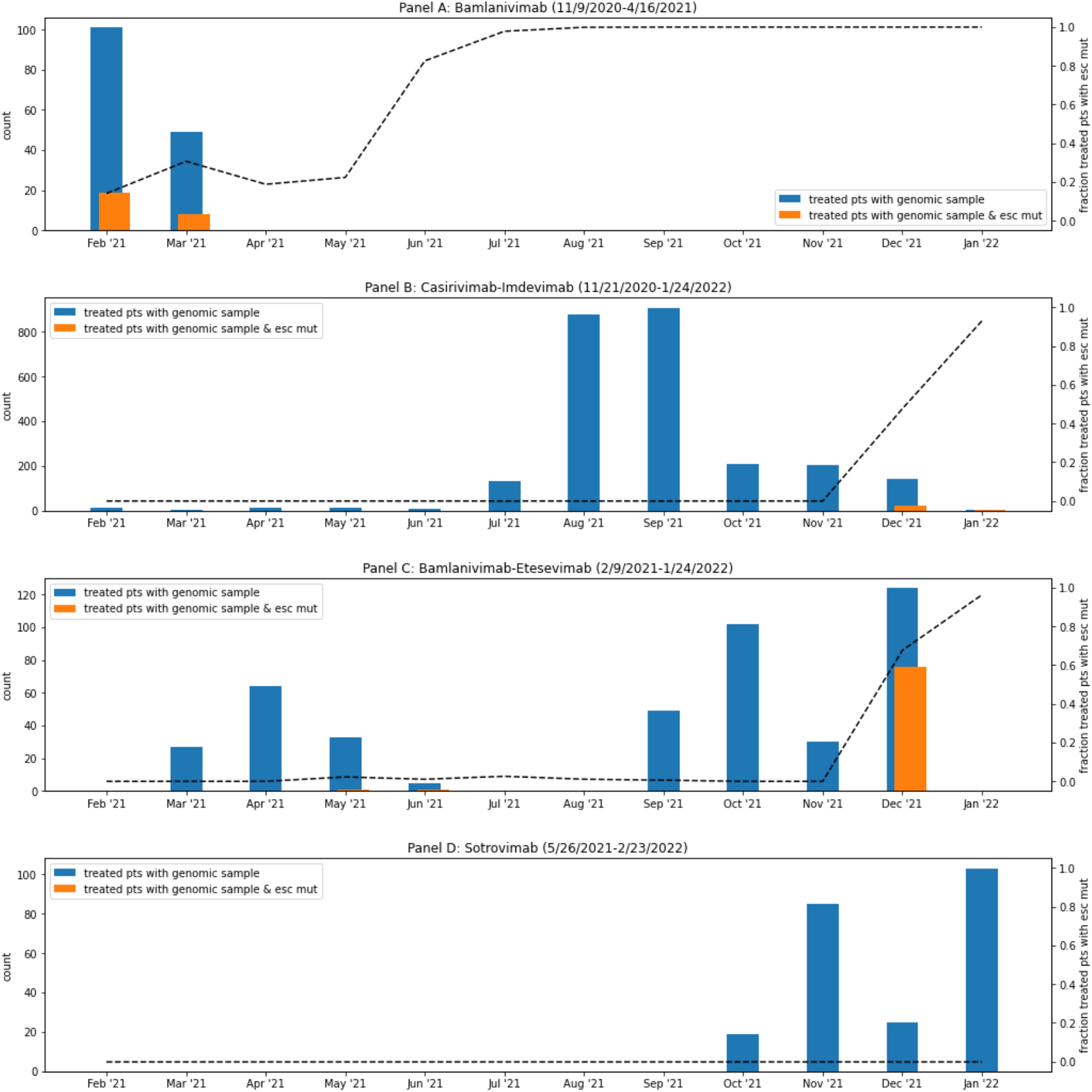
Number of Genomic Samples from nMAbs-Treated Patients with and without nMAb Escape Mutations. Number of sequenced samples from patients treated with bamlanivimab (Panel A), casirivimab + imdevimab (Panel B), bamlanivimab + etesevimab (Panel C), and sotrovimab (Panel D) by month of diagnosis (left axis). Blue bars: Total number of sequenced, treated samples. Orange bars: Number of sequenced samples with at least one putative escape mutation for each nMAb in the corresponding treatment. Dashed line: Fraction of all sequenced patient samples (treated and untreated) with at least one putative escape mutation to each nMAb in the corresponding treatment (scale on right axis). Dates of EUA for use of each nMAbs indicated above the graphs. Only putative escape mutations identified from deep mutational scanning experiments (i.e., those in Table 2) are included; results do not include S371F/L. Results exclude sequenced samples with missing escape mutation data and missing treatment status. Total numbers of samples considered across this timeframe (February 2021 to January 2022) are as follows: bamlanivimab, 150; casirivimab-imdevimab, 2,526; bamlanivimab-etesevimab, 434; sotrovimab, 232.

Figure 4 shows the distribution of sequence gaps and ambiguous base calls by position, relative to the reference genome, for three major WHO variants: Alpha, Delta, and Omicron. Although there are some overlaps (e.g., a relatively high frequency of samples from Alpha and Delta sharing gaps and ambiguous base calls near position 19500), there are notable differences across the variants. For example, in Omicron samples, gaps and ambiguous base calls are more common in the region of the genome encompassing the spike gene, relative to Alpha and Delta. Other distinct Omicron gap and ambiguous base call regions include those near positions 22700, 26300, and 27100.

**Figure 4.**
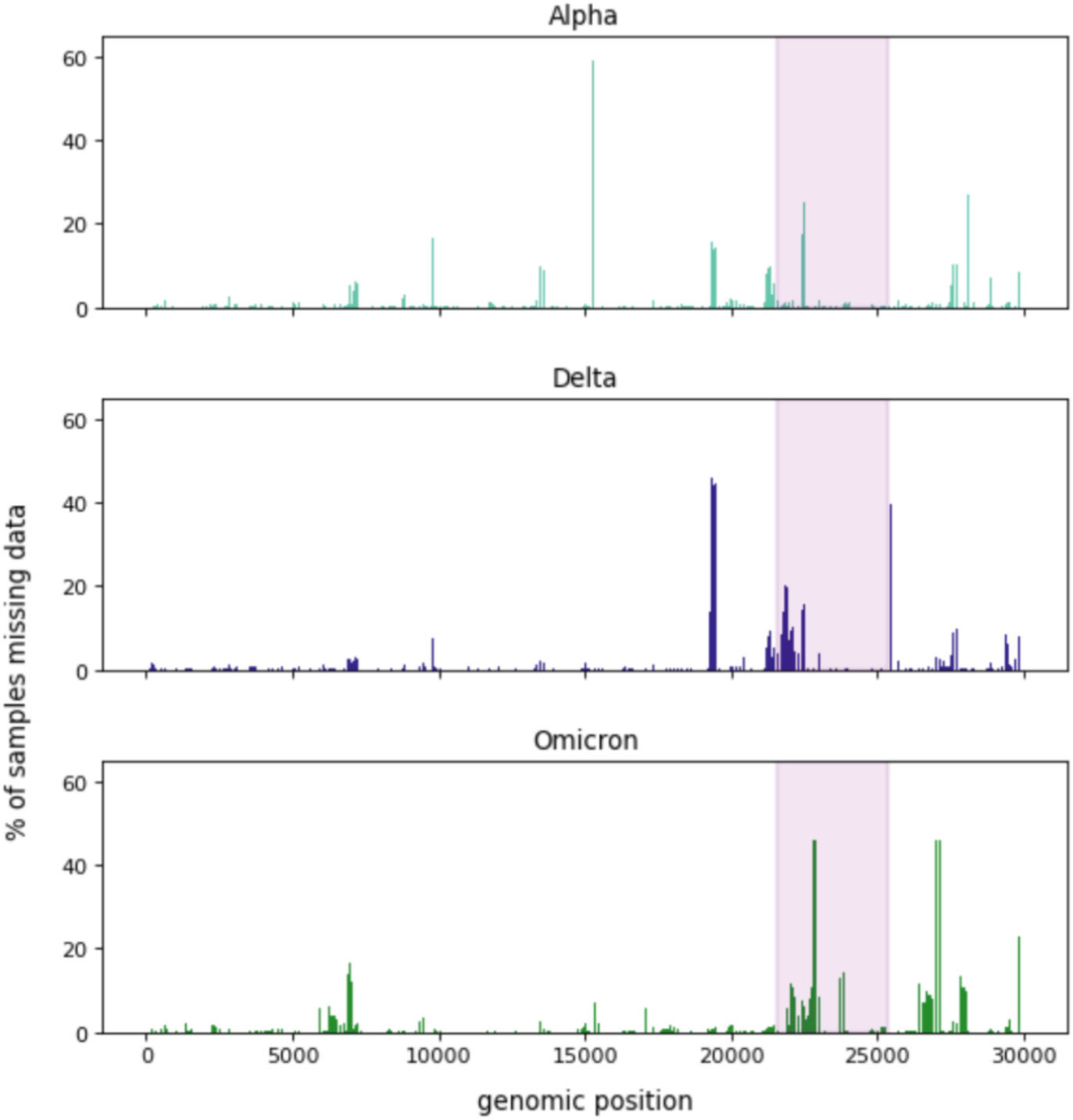
Frequency of Gaps and Ambiguous Base Calls by Genome Position (Relative to Reference Genome) The plots show for each genomic position relative to the reference genome the percentage of samples with apparent gaps or ambiguous base calls for Alpha, Delta, and Omicron variants. The shaded region represents the spike gene. All Alpha, Delta, and Omicron samples were considered for this analysis.

## CONCLUSIONS

This study presents a large, clinically derived data set of genomic sequences for SARS-CoV-2. The data from this study were part of a larger effort designed to assess the safety and effectiveness of nMAbs for treatment of COVID-19. This genomics data was combined with available clinical data within the centralized registry, which enabled investigation of clinical outcomes by confirmed SARS-CoV-2 variant in addition to the month and year of diagnosis and treatment status.^4^

The percentage of patients treated with nMAbs was higher in the group of patients with viral genomics data than in those without (30.2% vs. 12.6%). Aside from this, the ages, genders, racial and ethnic distribution, and distribution of several relevant comorbidities, along with smoking and pregnancy status, were similar in both groups. These observations are likely driven by two specific deliberate strategies used to obtain viral genomics data in the parent study: 1) maximize the number of nMAb-treated patients with viral genomics data, and 2) create a non-treated population with viral genomics data similar to the corresponding treated group of patients. Both strategies were adopted to facilitate nMAb effectiveness analysis across different WHO variants.

The distribution of WHO variants across epochs in the data set matches concurrent CDC genomic surveillance results across the United States.^14^ Large numbers of Delta and Omicron variant sequences within the data set reflect surges of infections observed across the pandemic, with respective viral variant dominance across June to November 2021 for Delta and across December 2021 to February 2022 for Omicron.

The genomics pipeline identified 13,522 unique mutations and the presence of 24 putative nMAb escape mutations previously shown to modify the epitope profile of the RBD of the S protein, resulting in interference of the binding of nMAbs in EUA-authorized treatments,^25^ including those in use during this study timeframe—bamlanivimab, etesevimab, imdevimab, casirivimab, sotrovimab, cilgavimab, and tixagevimab.^22,23^ Within the data set collected, at least one escape mutation was detected for each of these nMAbs and escape mutations within the RBD region increased in frequency with the emergence of Omicron. Timing of detection of escape mutations within clinical samples matched the timing of EUA amendment decisions.^26^ Interestingly, only 176 sequenced samples came from patients treated with an nMAb drug treatment for which their sample had putative escape mutations. In addition, the detection of mutations S371L and S371F in Omicron BA.1 and BA.2 samples is consistent with the timing of the U.S. Food and Drug Administration revoking the EUA for sotrovimab for treatment of COVID-19 in early April 2022.^27^ The small number of instances of patients harboring a virus with putative escape mutations to the nMAbs with which they were treated precluded further analysis along these lines. For instance, it would have been interesting to assess whether putative escape to a given treatment is associated with different clinical outcome risks; however, sample sizes did not support this type of analysis. This highlights the timeliness of FDA updates to EUA criteria for specific nMAbs.

This RWE study aggregated sequence data from four U.S. health systems. The high-quality sequences (≥80% of bases within reads with Phred score ≥30) were vetted to quality metrics to provide near complete genome coverage (≥85% coverage) at a depth that would allow confident base position calls (≥10 calls per genomic location). Setting these thresholds allowed confidence in the observations within the genomes and in the linkages to their respective clinical outcomes; however, the minimum metrics requirements increased data harmonization challenges across health systems. Standard metrics for assessing sequence quality, such as CT, were not universally collected and reported. Additionally, health systems, or associated sequencing centers, varied in protocols for library creation and timing of updates to protocols (e.g., ARTIC tiling PCR primers) to account for mutations within the viral population at large. Decisions about which samples to sequence were based on local factors, such as availability of resources and purpose of sequencing efforts beyond the research study. For example, a facility that wants to monitor only large-scale surveillance of the genomic population may not sequence at the same depth or breadth as a facility that wants to ask clinical impact questions of the genomes. As a result, the shallow read depths observed across a portion of the data resulted in their disqualification from the study.

Repurposing surveillance-quality data presents a significant challenge for genomics data harmonization for RWE studies such as this one. Further, the rapid mutation of the virus across the pandemic requires the periodic update of sequencing protocols and primer sets to address regions that may be dropped due to mutation accumulations, as was observed in the emergent Omicron variant.^28^ Local processes are unable to be completely controlled and normalized across multiple sequencing centers. For the future, continued monitoring of minimum sequence quality thresholds would serve to identify sequencing challenges (e.g., primer dropout due to new variant mutations) and to detect newly emerging variants sooner.

The novelty of the parent real-world evidence (RWE) study of the effectiveness of nMAbs is the use of viral genome sequence data with clinical data to make impactful, clinically relevant observations.^4^ The assignment of samples to WHO nomenclature using genomics data enabled analyses of health outcomes and nMAbs effectiveness by WHO variant in U.S. patients over time,^4^ which was essential during times when no single variant was dominant. Further analyses of nucleotide variants provided information about potential nMAb escape mutations. Continuation of real-world data collection that connects real-time surveillance of linked genomic and clinical data could help to identify clinically significant mutations and better prepare for new variants prior to lineage definition. Having navigated the challenges in sharing and combining data across geographically distributed health systems, this study represents a further step in developing a community network for more effective surveillance.^29^

## Data Availability

Deidentified data has been submitted to National COVID Cohort Collaborative (N3C). Available at https://ncats.nih.gov/n3c.

## ACKNOWLEDGMENTS

This work was performed by the mAb Real World Evidence Collaborative (authors listed alphabetically). The views expressed are solely those of the authors and do not necessarily represent those of the U.S. Department of Health and Human Services. This study was supported wholly or in part with federal funds from the Administration for Strategic Preparedness and Response, Biomedical Advanced Research and Development Authority, under Contract Number 75FCMC18D0047, Task Order 75A50121F80012 awarded to The MITRE Corporation.

## NOTICE

This (software/technical data) was produced for the U. S. Government under Contract Number 75FCMC18D0047, and is subject to Federal Acquisition Regulation Clause 52.227-14, Rights in Data-General.

No other use other than that granted to the U. S. Government, or to those acting on behalf of the U. S. Government under that Clause is authorized without the express written permission of The MITRE Corporation.

For further information, please contact The MITRE Corporation, Contracts Management Office, 7515 Colshire Drive, McLean, VA 22102-7539, (703) 983-6000.

