## Supplementary Material for "Descriptive Analysis of SARS-CoV-2 Genomics Data from Ambulatory Patients"

Approved for Public Release; Distribution Unlimited. Public Release Case Number 22-2693
© 2023 The MITRE Corporation. All Rights Reserved.

Table S1. Complete List of PANGO Lineage Calls and WHO Variant Equivalents for Viral Genomic Samples within Study

The total number of samples with a PANGO lineage call is 14,796; the number of samples with an PANGO lineage call and a definitive nMAb treatment status is 13,703.

| PANGO Lineage | WHO Variant | Total (n) | Count in nMAbs Effectiveness Study Cohort | Count Non-Treated | Count Treated |
|---|---|---|---|---|---|
| A.2.5 | no_WHO_equivalent | 1 | 1 | 1 | 0 |
| AY.1 | Delta | 5 | 4 | 4 | 0 |
| AY.100 | Delta | 743 | 733 | 368 | 365 |
| AY.103 | Delta | 1,499 | 1,412 | 788 | 624 |
| AY.105 | Delta | 6 | 5 | 2 | 3 |
| AY.107 | Delta | 35 | 35 | 27 | 8 |
| AY.109 | Delta | 3 | 3 | 2 | 1 |
| AY.110 | Delta | 9 | 8 | 4 | 4 |
| AY.112 | Delta | 104 | 101 | 84 | 17 |
| AY.113 | Delta | 39 | 37 | 18 | 19 |
| AY.114 | Delta | 20 | 19 | 7 | 12 |
| AY.116 | Delta | 1 | 1 | 1 | 0 |
| AY.116.1 | Delta | 8 | 7 | 3 | 4 |
| AY.117 | Delta | 46 | 44 | 17 | 27 |
| AY.118 | Delta | 58 | 51 | 28 | 23 |
| AY.119 | Delta | 80 | 75 | 44 | 31 |
| AY.119.2 | Delta | 4 | 4 | 2 | 2 |
| AY.120 | Delta | 59 | 56 | 51 | 5 |
| AY.120.1 | Delta | 10 | 10 | 8 | 2 |
| AY.121 | Delta | 3 | 2 | 0 | 2 |
| AY.122 | Delta | 74 | 74 | 48 | 26 |
| AY.125 | Delta | 1 | 1 | 0 | 1 |
| AY.126 | Delta | 2 | 2 | 2 | 0 |
| AY.127 | Delta | 5 | 4 | 4 | 0 |

Approved for Public Release; Distribution Unlimited. Public Release Case Number 22-2693
© 2023 The MITRE Corporation. All Rights Reserved.

| PANGO Lineage | WHO Variant | Total (n) | Count in nMAbs Effectiveness Study Cohort | Count Non-Treated | Count Treated |
|---|---|---|---|---|---|
| AY.129 | Delta | 18 | 18 | 9 | 9 |
| AY.13 | Delta | 22 | 21 | 12 | 9 |
| AY.14 | Delta | 18 | 15 | 14 | 1 |
| AY.15 | Delta | 9 | 9 | 7 | 2 |
| AY.16 | Delta | 8 | 6 | 6 | 0 |
| AY.2 | Delta | 18 | 17 | 14 | 3 |
| AY.20 | Delta | 91 | 88 | 62 | 26 |
| AY.23 | Delta | 6 | 5 | 4 | 1 |
| AY.25 | Delta | 1,074 | 1,046 | 606 | 440 |
| AY.25.1 | Delta | 199 | 185 | 95 | 90 |
| AY.25.2 | Delta | 1 | 1 | 0 | 1 |
| AY.25.3 | Delta | 14 | 13 | 8 | 5 |
| AY.26 | Delta | 188 | 180 | 118 | 62 |
| AY.3 | Delta | 721 | 695 | 352 | 343 |
| AY.3.1 | Delta | 60 | 60 | 30 | 30 |
| AY.3.3 | Delta | 1 | 1 | 1 | 0 |
| AY.30 | Delta | 1 | 1 | 1 | 0 |
| AY.34 | Delta | 2 | 2 | 1 | 1 |
| AY.34.1 | Delta | 4 | 4 | 1 | 3 |
| AY.35 | Delta | 17 | 16 | 9 | 7 |
| AY.36 | Delta | 4 | 3 | 3 | 0 |
| AY.37 | Delta | 11 | 10 | 3 | 7 |
| AY.39 | Delta | 173 | 163 | 75 | 88 |
| AY.39.1 | Delta | 9 | 9 | 3 | 6 |
| AY.4 | Delta | 33 | 33 | 20 | 13 |
| AY.4.6 | Delta | 1 | 1 | 0 | 1 |
| AY.4.7 | Delta | 5 | 5 | 5 | 0 |

Approved for Public Release; Distribution Unlimited. Public Release Case Number 22-2693
© 2023 The MITRE Corporation. All Rights Reserved.

| PANGO Lineage | WHO Variant | Total (n) | Count in nMAbs Effectiveness Study Cohort | Count Non-Treated | Count Treated |
|---|---|---|---|---|---|
| AY.42 | Delta | 7 | 7 | 5 | 2 |
| AY.43 | Delta | 18 | 16 | 15 | 1 |
| AY.44 | Delta | 2,576 | 2,485 | 1,918 | 567 |
| AY.46 | Delta | 8 | 6 | 4 | 2 |
| AY.46.4 | Delta | 41 | 39 | 19 | 20 |
| AY.47 | Delta | 219 | 209 | 110 | 99 |
| AY.48 | Delta | 13 | 13 | 12 | 1 |
| AY.49 | Delta | 1 | 1 | 0 | 1 |
| AY.5 | Delta | 1 | 1 | 0 | 1 |
| AY.5.3 | Delta | 2 | 2 | 0 | 2 |
| AY.52 | Delta | 7 | 7 | 7 | 0 |
| AY.54 | Delta | 188 | 181 | 124 | 57 |
| AY.59 | Delta | 6 | 6 | 5 | 1 |
| AY.62 | Delta | 8 | 8 | 4 | 4 |
| AY.64 | Delta | 16 | 16 | 6 | 10 |
| AY.67 | Delta | 11 | 10 | 6 | 4 |
| AY.7.1 | Delta | 1 | 1 | 0 | 1 |
| AY.74 | Delta | 5 | 5 | 2 | 3 |
| AY.75 | Delta | 92 | 90 | 44 | 46 |
| AY.77 | Delta | 7 | 7 | 5 | 2 |
| AY.80 | Delta | 1 | 1 | 1 | 0 |
| AY.81 | Delta | 34 | 34 | 30 | 4 |
| AY.82 | Delta | 1 | 1 | 1 | 0 |
| AY.83 | Delta | 6 | 4 | 2 | 2 |
| AY.86 | Delta | 2 | 2 | 1 | 1 |
| AY.88 | Delta | 2 | 2 | 1 | 1 |
| AY.9.2 | Delta | 12 | 10 | 5 | 5 |

Approved for Public Release; Distribution Unlimited. Public Release Case Number 22-2693
© 2023 The MITRE Corporation. All Rights Reserved.

| PANGO Lineage | WHO Variant | Total (n) | Count in nMAbs Effectiveness Study Cohort | Count Non-Treated | Count Treated |
|---|---|---|---|---|---|
| AY.91 | Delta | 1 | 1 | 0 | 1 |
| AY.92 | Delta | 1 | 1 | 1 | 0 |
| AY.98.1 | Delta | 8 | 8 | 4 | 4 |
| AZ.3 | no_WHO_equivalent | 4 | 4 | 4 | 0 |
| B | no_WHO_equivalent | 4 | 3 | 3 | 0 |
| B.1 | no_WHO_equivalent | 154 | 146 | 121 | 25 |
| B.1.1 | no_WHO_equivalent | 10 | 9 | 6 | 3 |
| B.1.1.186 | no_WHO_equivalent | 1 | 1 | 1 | 0 |
| B.1.1.192 | no_WHO_equivalent | 1 | 1 | 1 | 0 |
| B.1.1.222 | no_WHO_equivalent | 30 | 27 | 17 | 10 |
| B.1.1.239 | no_WHO_equivalent | 1 | 1 | 1 | 0 |
| B.1.1.265 | no_WHO_equivalent | 1 | 0 | 0 | 0 |
| B.1.1.318 | no_WHO_equivalent | 9 | 9 | 9 | 0 |
| B.1.1.416 | no_WHO_equivalent | 12 | 10 | 4 | 6 |
| B.1.1.432 | no_WHO_equivalent | 3 | 3 | 1 | 2 |
| B.1.1.519 | no_WHO_equivalent | 127 | 119 | 102 | 17 |
| B.1.1.7 | Alpha | 1,082 | 1061 | 857 | 204 |
| B.1.1.8 | no_WHO_equivalent | 1 | 0 | 0 | 0 |
| B.1.110.3 | no_WHO_equivalent | 1 | 0 | 0 | 0 |
| B.1.126 | no_WHO_equivalent | 1 | 1 | 1 | 0 |
| B.1.2 | no_WHO_equivalent | 360 | 343 | 227 | 116 |
| B.1.222 | no_WHO_equivalent | 1 | 1 | 1 | 0 |
| B.1.232 | no_WHO_equivalent | 3 | 2 | 2 | 0 |
| B.1.234 | no_WHO_equivalent | 13 | 11 | 5 | 6 |
| B.1.239 | no_WHO_equivalent | 4 | 4 | 4 | 0 |
| B.1.240 | no_WHO_equivalent | 2 | 2 | 1 | 1 |
| B.1.241 | no_WHO_equivalent | 2 | 2 | 2 | 0 |

Approved for Public Release; Distribution Unlimited. Public Release Case Number 22-2693
© 2023 The MITRE Corporation. All Rights Reserved.

| PANGO Lineage | WHO Variant | Total (n) | Count in nMAbs Effectiveness Study Cohort | Count Non-Treated | Count Treated |
|---|---|---|---|---|---|
| B.1.243 | no_WHO_equivalent | 12 | 12 | 6 | 6 |
| B.1.258 | no_WHO_equivalent | 2 | 2 | 2 | 0 |
| B.1.258.23 | no_WHO_equivalent | 8 | 8 | 7 | 1 |
| B.1.265 | no_WHO_equivalent | 1 | 1 | 1 | 0 |
| B.1.313 | no_WHO_equivalent | 4 | 4 | 4 | 0 |
| B.1.324 | no_WHO_equivalent | 1 | 1 | 1 | 0 |
| B.1.349 | no_WHO_equivalent | 1 | 1 | 1 | 0 |
| B.1.351 | Beta | 9 | 9 | 8 | 1 |
| B.1.36.7 | no_WHO_equivalent | 1 | 1 | 1 | 0 |
| B.1.369 | no_WHO_equivalent | 2 | 2 | 2 | 0 |
| B.1.375 | no_WHO_equivalent | 1 | 1 | 1 | 0 |
| B.1.396 | no_WHO_equivalent | 2 | 2 | 1 | 1 |
| B.1.399 | no_WHO_equivalent | 1 | 0 | 0 | 0 |
| B.1.400 | no_WHO_equivalent | 16 | 16 | 14 | 2 |
| B.1.404 | no_WHO_equivalent | 2 | 2 | 2 | 0 |
| B.1.424 | no_WHO_equivalent | 1 | 1 | 0 | 1 |
| B.1.426 | no_WHO_equivalent | 1 | 1 | 1 | 0 |
| B.1.427 | Epsilon | 67 | 65 | 52 | 13 |
| B.1.429 | Epsilon | 284 | 237 | 165 | 72 |
| B.1.433 | no_WHO_equivalent | 5 | 2 | 0 | 2 |
| B.1.525 | Eta | 2 | 2 | 0 | 2 |
| B.1.526 | Iota | 19 | 18 | 16 | 2 |
| B.1.544 | no_WHO_equivalent | 1 | 1 | 1 | 0 |
| B.1.551 | no_WHO_equivalent | 3 | 3 | 1 | 2 |
| B.1.561 | no_WHO_equivalent | 23 | 20 | 15 | 5 |
| B.1.565 | no_WHO_equivalent | 1 | 1 | 1 | 0 |
| B.1.568 | no_WHO_equivalent | 1 | 1 | 0 | 1 |

Approved for Public Release; Distribution Unlimited. Public Release Case Number 22-2693
© 2023 The MITRE Corporation. All Rights Reserved.

| PANGO Lineage | WHO Variant | Total (n) | Count in nMAbs Effectiveness Study Cohort | Count Non-Treated | Count Treated |
|---|---|---|---|---|---|
| B.1.575 | no_WHO_equivalent | 15 | 15 | 15 | 0 |
| B.1.577 | no_WHO_equivalent | 1 | 1 | 1 | 0 |
| B.1.582 | no_WHO_equivalent | 1 | 1 | 0 | 1 |
| B.1.587 | no_WHO_equivalent | 2 | 2 | 2 | 0 |
| B.1.595 | no_WHO_equivalent | 12 | 9 | 7 | 2 |
| B.1.596 | no_WHO_equivalent | 3 | 3 | 3 | 0 |
| B.1.599 | no_WHO_equivalent | 1 | 1 | 1 | 0 |
| B.1.609 | no_WHO_equivalent | 8 | 6 | 4 | 2 |
| B.1.612 | no_WHO_equivalent | 3 | 3 | 3 | 0 |
| B.1.617.1 | Kappa | 1 | 1 | 1 | 0 |
| B.1.617.2 | Delta | 241 | 234 | 148 | 86 |
| B.1.621 | Mu | 66 | 66 | 59 | 7 |
| B.1.621.1 | Mu | 1 | 1 | 1 | 0 |
| B.1.621.2 | Mu | 1 | 1 | 1 | 0 |
| B.1.627 | no_WHO_equivalent | 2 | 2 | 1 | 1 |
| B.1.631 | no_WHO_equivalent | 1 | 1 | 0 | 1 |
| B.1.632 | no_WHO_equivalent | 2 | 2 | 1 | 1 |
| B.1.637 | no_WHO_equivalent | 7 | 7 | 3 | 4 |
| BA.1 | Omicron | 46 | 43 | 33 | 10 |
| BA.1.1 | Omicron | 1,484 | 1,131 | 1,011 | 120 |
| BA.1.1.1 | Omicron | 30 | 28 | 25 | 3 |
| BA.1.1.10 | Omicron | 3 | 3 | 2 | 1 |
| BA.1.1.11 | Omicron | 31 | 25 | 21 | 4 |
| BA.1.1.12 | Omicron | 1 | 0 | 0 | 0 |
| BA.1.1.13 | Omicron | 1 | 1 | 1 | 0 |
| BA.1.1.14 | Omicron | 22 | 18 | 15 | 3 |
| BA.1.1.15 | Omicron | 10 | 10 | 9 | 1 |

Approved for Public Release; Distribution Unlimited. Public Release Case Number 22-2693
© 2023 The MITRE Corporation. All Rights Reserved.

| PANGO Lineage | WHO Variant | Total (n) | Count in nMAbs Effectiveness Study Cohort | Count Non-Treated | Count Treated |
|---|---|---|---|---|---|
| BA.1.1.16 | Omicron | 15 | 11 | 9 | 2 |
| BA.1.1.2 | Omicron | 18 | 15 | 15 | 0 |
| BA.1.1.4 | Omicron | 1 | 1 | 1 | 0 |
| BA.1.1.5 | Omicron | 1 | 1 | 1 | 0 |
| BA.1.1.8 | Omicron | 3 | 3 | 3 | 0 |
| BA.1.13 | Omicron | 8 | 7 | 7 | 0 |
| BA.1.14 | Omicron | 183 | 144 | 134 | 10 |
| BA.1.15 | Omicron | 903 | 832 | 558 | 274 |
| BA.1.15.1 | Omicron | 36 | 35 | 33 | 2 |
| BA.1.15.2 | Omicron | 6 | 6 | 2 | 4 |
| BA.1.16 | Omicron | 42 | 40 | 38 | 2 |
| BA.1.17 | Omicron | 14 | 13 | 12 | 1 |
| BA.1.17.2 | Omicron | 20 | 20 | 19 | 1 |
| BA.1.18 | Omicron | 26 | 23 | 18 | 5 |
| BA.1.19 | Omicron | 9 | 6 | 5 | 1 |
| BA.1.20 | Omicron | 251 | 153 | 122 | 31 |
| BA.1.21 | Omicron | 3 | 3 | 2 | 1 |
| BA.1.5 | Omicron | 2 | 2 | 2 | 0 |
| BA.1.6 | Omicron | 1 | 0 | 0 | 0 |
| BA.1.9 | Omicron | 5 | 5 | 4 | 1 |
| BA.2 | Omicron | 2 | 1 | 1 | 0 |
| BA.2.3 | Omicron | 3 | 1 | 1 | 0 |
| BA.2.5 | Omicron | 4 | 1 | 1 | 0 |
| BA.2.9 | Omicron | 3 | 0 | 0 | 0 |
| C.36 | no_WHO_equivalent | 1 | 1 | 1 | 0 |
| C.37 | Lambda | 4 | 4 | 4 | 0 |
| P.1 | Gamma | 71 | 69 | 61 | 8 |

Approved for Public Release; Distribution Unlimited. Public Release Case Number 22-2693
© 2023 The MITRE Corporation. All Rights Reserved.

| PANGO Lineage | WHO Variant | Total<br><br>(n) | Count in nMAbs Effectiveness Study Cohort | Count Non-Treated | Count Treated |
|---|---|---|---|---|---|
| P.1.1 | Gamma | 3 | 3 | 3 | 0 |
| P.1.10 | Gamma | 3 | 3 | 3 | 0 |
| P.1.14 | Gamma | 2 | 2 | 2 | 0 |
| P.1.15 | Gamma | 1 | 1 | 1 | 0 |
| P.1.17 | Gamma | 2 | 2 | 2 | 0 |
| P.2 | Zeta | 9 | 8 | 6 | 2 |
| Q.3 | Alpha | 24 | 23 | 22 | 1 |
| Q.8 | Alpha | 1 | 1 | 0 | 1 |
| R.1 | no_WHO_equivalent | 2 | 2 | 1 | 1 |
| XB | no_WHO_equivalent | 2 | 2 | 1 | 1 |

Approved for Public Release; Distribution Unlimited. Public Release Case Number 22-2693
© 2023 The MITRE Corporation. All Rights Reserved.

**Table S2. Number of Sequences by PANGO Lineage and Diagnosis Epoch**

The total number of samples is 14,796.

| PANGO Lineage | 2020 | | | | | | 2021 | | | | | | | | | | | | 2022 | | |
|---|---|---|---|---|---|---|---|---|---|---|---|---|---|---|---|---|---|---|---|---|---|
| | July | Aug | Sep | Oct | Nov | Dec | Jan | Feb | Mar | Apr | May | June | July | Aug | Sep | Oct | Nov | Dec | Jan | Feb | Mar |
| A.2.5 | 0 | 0 | 0 | 0 | 0 | 0 | 0 | 0 | 0 | 1 | 0 | 0 | 0 | 0 | 0 | 0 | 0 | 0 | 0 | 0 | 0 |
| AY.1 | 0 | 0 | 0 | 0 | 0 | 0 | 0 | 0 | 0 | 0 | 0 | 0 | 0 | 2 | 2 | 0 | 0 | 1 | 0 | 0 | 0 |
| AY.100 | 0 | 0 | 0 | 0 | 0 | 0 | 0 | 0 | 0 | 0 | 0 | 0 | 26 | 271 | 243 | 79 | 70 | 42 | 12 | 0 | 0 |
| AY.103 | 0 | 0 | 0 | 0 | 0 | 0 | 0 | 0 | 0 | 0 | 0 | 8 | 55 | 379 | 507 | 169 | 234 | 120 | 27 | 0 | 0 |
| AY.105 | 0 | 0 | 0 | 0 | 0 | 0 | 0 | 0 | 0 | 0 | 0 | 1 | 2 | 2 | 1 | 0 | 0 | 0 | 0 | 0 | 0 |
| AY.107 | 0 | 0 | 0 | 0 | 0 | 0 | 0 | 0 | 0 | 0 | 0 | 0 | 0 | 4 | 4 | 2 | 13 | 9 | 3 | 0 | 0 |
| AY.109 | 0 | 0 | 0 | 0 | 0 | 0 | 0 | 0 | 0 | 0 | 0 | 0 | 0 | 0 | 2 | 0 | 1 | 0 | 0 | 0 | 0 |
| AY.110 | 0 | 0 | 0 | 0 | 0 | 0 | 0 | 0 | 0 | 0 | 0 | 0 | 3 | 6 | 0 | 0 | 0 | 0 | 0 | 0 | 0 |
| AY.112 | 0 | 0 | 0 | 0 | 0 | 0 | 0 | 0 | 0 | 0 | 0 | 2 | 4 | 25 | 46 | 10 | 12 | 5 | 0 | 0 | 0 |
| AY.113 | 0 | 0 | 0 | 0 | 0 | 0 | 0 | 0 | 0 | 0 | 0 | 0 | 2 | 10 | 12 | 4 | 5 | 4 | 2 | 0 | 0 |
| AY.114 | 0 | 0 | 0 | 0 | 0 | 0 | 0 | 0 | 0 | 0 | 0 | 0 | 0 | 7 | 8 | 3 | 1 | 1 | 0 | 0 | 0 |
| AY.116 | 0 | 0 | 0 | 0 | 0 | 0 | 0 | 0 | 0 | 0 | 0 | 0 | 0 | 0 | 1 | 0 | 0 | 0 | 0 | 0 | 0 |
| AY.116.1 | 0 | 0 | 0 | 0 | 0 | 0 | 0 | 0 | 0 | 0 | 0 | 0 | 1 | 1 | 5 | 0 | 0 | 1 | 0 | 0 | 0 |
| AY.117 | 0 | 0 | 0 | 0 | 0 | 0 | 0 | 0 | 0 | 0 | 0 | 0 | 3 | 10 | 15 | 1 | 7 | 10 | 0 | 0 | 0 |
| AY.118 | 0 | 0 | 0 | 0 | 0 | 0 | 0 | 0 | 0 | 0 | 0 | 0 | 5 | 22 | 20 | 8 | 2 | 1 | 0 | 0 | 0 |
| AY.119 | 0 | 0 | 0 | 0 | 0 | 0 | 0 | 0 | 0 | 0 | 0 | 0 | 9 | 26 | 15 | 2 | 13 | 11 | 4 | 0 | 0 |
| AY.119.2 | 0 | 0 | 0 | 0 | 0 | 0 | 0 | 0 | 0 | 0 | 0 | 0 | 0 | 0 | 2 | 1 | 1 | 0 | 0 | 0 | 0 |

Approved for Public Release; Distribution Unlimited. Public Release Case Number 22-2693
© 2023 The MITRE Corporation. All Rights Reserved.

| PANGO Lineage | 2020 | | | | | | 2021 | | | | | | | | | | | | 2022 | | |
|---|---|---|---|---|---|---|---|---|---|---|---|---|---|---|---|---|---|---|---|---|---|
| | July | Aug | Sep | Oct | Nov | Dec | Jan | Feb | Mar | Apr | May | June | July | Aug | Sep | Oct | Nov | Dec | Jan | Feb | Mar |
| AY.120 | 0 | 0 | 0 | 0 | 0 | 0 | 0 | 0 | 0 | 0 | 0 | 0 | 1 | 16 | 30 | 6 | 5 | 1 | 0 | 0 | 0 |
| AY.120.1 | 0 | 0 | 0 | 0 | 0 | 0 | 0 | 0 | 0 | 0 | 0 | 0 | 1 | 3 | 5 | 1 | 0 | 0 | 0 | 0 | 0 |
| AY.121 | 0 | 0 | 0 | 0 | 0 | 0 | 0 | 0 | 0 | 0 | 0 | 0 | 0 | 0 | 1 | 0 | 2 | 0 | 0 | 0 | 0 |
| AY.122 | 0 | 0 | 0 | 0 | 0 | 0 | 0 | 0 | 0 | 0 | 1 | 4 | 8 | 14 | 25 | 8 | 5 | 9 | 0 | 0 | 0 |
| AY.125 | 0 | 0 | 0 | 0 | 0 | 0 | 0 | 0 | 0 | 0 | 0 | 0 | 0 | 1 | 0 | 0 | 0 | 0 | 0 | 0 | 0 |
| AY.126 | 0 | 0 | 0 | 0 | 0 | 0 | 0 | 0 | 0 | 0 | 0 | 0 | 0 | 1 | 0 | 0 | 1 | 0 | 0 | 0 | 0 |
| AY.127 | 0 | 0 | 0 | 0 | 0 | 0 | 0 | 0 | 0 | 0 | 0 | 0 | 0 | 1 | 0 | 0 | 1 | 3 | 0 | 0 | 0 |
| AY.129 | 0 | 0 | 0 | 0 | 0 | 0 | 0 | 0 | 0 | 0 | 0 | 0 | 0 | 0 | 7 | 1 | 10 | 0 | 0 | 0 | 0 |
| AY.13 | 0 | 0 | 0 | 0 | 0 | 0 | 0 | 0 | 0 | 0 | 0 | 0 | 4 | 12 | 5 | 1 | 0 | 0 | 0 | 0 | 0 |
| AY.14 | 0 | 0 | 0 | 0 | 0 | 0 | 0 | 0 | 0 | 0 | 0 | 3 | 5 | 7 | 3 | 0 | 0 | 0 | 0 | 0 | 0 |
| AY.15 | 0 | 0 | 0 | 0 | 0 | 0 | 0 | 0 | 0 | 0 | 0 | 3 | 1 | 5 | 0 | 0 | 0 | 0 | 0 | 0 | 0 |
| AY.16 | 0 | 0 | 0 | 0 | 0 | 0 | 0 | 0 | 0 | 0 | 0 | 0 | 0 | 2 | 6 | 0 | 0 | 0 | 0 | 0 | 0 |
| AY.2 | 0 | 0 | 0 | 0 | 0 | 0 | 0 | 0 | 0 | 0 | 0 | 0 | 3 | 11 | 4 | 0 | 0 | 0 | 0 | 0 | 0 |
| AY.20 | 0 | 0 | 0 | 0 | 0 | 0 | 0 | 0 | 0 | 0 | 0 | 1 | 6 | 43 | 14 | 14 | 6 | 7 | 0 | 0 | 0 |
| AY.23 | 0 | 0 | 0 | 0 | 0 | 0 | 0 | 0 | 0 | 0 | 0 | 0 | 1 | 1 | 3 | 0 | 0 | 0 | 1 | 0 | 0 |
| AY.25 | 0 | 0 | 0 | 0 | 0 | 0 | 0 | 0 | 0 | 0 | 0 | 11 | 46 | 383 | 331 | 111 | 131 | 54 | 7 | 0 | 0 |
| AY.25.1 | 0 | 0 | 0 | 0 | 0 | 0 | 0 | 0 | 0 | 0 | 0 | 1 | 10 | 54 | 57 | 21 | 28 | 21 | 7 | 0 | 0 |
| AY.25.2 | 0 | 0 | 0 | 0 | 0 | 0 | 0 | 0 | 0 | 0 | 0 | 0 | 0 | 1 | 0 | 0 | 0 | 0 | 0 | 0 | 0 |
| AY.25.3 | 0 | 0 | 0 | 0 | 0 | 0 | 0 | 0 | 0 | 0 | 0 | 0 | 0 | 2 | 3 | 3 | 3 | 3 | 0 | 0 | 0 |

Approved for Public Release; Distribution Unlimited. Public Release Case Number 22-2693
© 2023 The MITRE Corporation. All Rights Reserved.

| PANGO Lineage | 2020 | | | | | | 2021 | | | | | | | | | | | | 2022 | | |
|---|---|---|---|---|---|---|---|---|---|---|---|---|---|---|---|---|---|---|---|---|---|
| | July | Aug | Sep | Oct | Nov | Dec | Jan | Feb | Mar | Apr | May | June | July | Aug | Sep | Oct | Nov | Dec | Jan | Feb | Mar |
| AY.26 | 0 | 0 | 0 | 0 | 0 | 0 | 0 | 0 | 0 | 0 | 0 | 2 | 20 | 75 | 51 | 13 | 17 | 9 | 1 | 0 | 0 |
| AY.3 | 0 | 0 | 0 | 0 | 0 | 0 | 0 | 0 | 0 | 0 | 0 | 1 | 35 | 201 | 234 | 75 | 110 | 55 | 10 | 0 | 0 |
| AY.3.1 | 0 | 0 | 0 | 0 | 0 | 0 | 0 | 0 | 0 | 0 | 0 | 0 | 4 | 29 | 20 | 3 | 4 | 0 | 0 | 0 | 0 |
| AY.3.3 | 0 | 0 | 0 | 0 | 0 | 0 | 0 | 0 | 0 | 0 | 0 | 0 | 0 | 0 | 0 | 0 | 0 | 0 | 1 | 0 | 0 |
| AY.30 | 0 | 0 | 0 | 0 | 0 | 0 | 0 | 0 | 0 | 0 | 0 | 0 | 0 | 1 | 0 | 0 | 0 | 0 | 0 | 0 | 0 |
| AY.34 | 0 | 0 | 0 | 0 | 0 | 0 | 0 | 0 | 0 | 0 | 0 | 0 | 0 | 0 | 0 | 0 | 1 | 1 | 0 | 0 | 0 |
| AY.34.1 | 0 | 0 | 0 | 0 | 0 | 0 | 0 | 0 | 0 | 0 | 0 | 0 | 0 | 2 | 0 | 0 | 0 | 2 | 0 | 0 | 0 |
| AY.35 | 0 | 0 | 0 | 0 | 0 | 0 | 0 | 0 | 0 | 0 | 0 | 2 | 4 | 8 | 3 | 0 | 0 | 0 | 0 | 0 | 0 |
| AY.36 | 0 | 0 | 0 | 0 | 0 | 0 | 0 | 0 | 0 | 0 | 0 | 0 | 0 | 0 | 3 | 0 | 1 | 0 | 0 | 0 | 0 |
| AY.37 | 0 | 0 | 0 | 0 | 0 | 0 | 0 | 0 | 0 | 0 | 0 | 0 | 1 | 2 | 8 | 0 | 0 | 0 | 0 | 0 | 0 |
| AY.39 | 0 | 0 | 0 | 0 | 0 | 0 | 0 | 0 | 0 | 0 | 0 | 0 | 1 | 23 | 42 | 23 | 60 | 19 | 5 | 0 | 0 |
| AY.39.1 | 0 | 0 | 0 | 0 | 0 | 0 | 0 | 0 | 0 | 0 | 0 | 0 | 0 | 2 | 5 | 0 | 1 | 1 | 0 | 0 | 0 |
| AY.4 | 0 | 0 | 0 | 0 | 0 | 0 | 0 | 0 | 0 | 0 | 0 | 0 | 1 | 3 | 12 | 7 | 5 | 5 | 0 | 0 | 0 |
| AY.4.6 | 0 | 0 | 0 | 0 | 0 | 0 | 0 | 0 | 0 | 0 | 0 | 0 | 0 | 0 | 1 | 0 | 0 | 0 | 0 | 0 | 0 |
| AY.4.7 | 0 | 0 | 0 | 0 | 0 | 0 | 0 | 0 | 0 | 0 | 0 | 0 | 0 | 1 | 2 | 2 | 0 | 0 | 0 | 0 | 0 |
| AY.42 | 0 | 0 | 0 | 0 | 0 | 0 | 0 | 0 | 0 | 0 | 0 | 0 | 0 | 3 | 4 | 0 | 0 | 0 | 0 | 0 | 0 |
| AY.43 | 0 | 0 | 0 | 0 | 0 | 0 | 0 | 0 | 0 | 0 | 0 | 0 | 1 | 3 | 7 | 0 | 4 | 2 | 1 | 0 | 0 |
| AY.44 | 0 | 0 | 0 | 0 | 0 | 0 | 0 | 0 | 0 | 2 | 33 | 314 | 358 | 773 | 638 | 164 | 182 | 92 | 20 | 0 | 0 |
| AY.46 | 0 | 0 | 0 | 0 | 0 | 0 | 0 | 0 | 0 | 0 | 0 | 0 | 0 | 2 | 3 | 2 | 1 | 0 | 0 | 0 | 0 |

Approved for Public Release; Distribution Unlimited. Public Release Case Number 22-2693
© 2023 The MITRE Corporation. All Rights Reserved.

| PANGO Lineage | 2020 | | | | | | 2021 | | | | | | | | | | | | 2022 | | |
|---|---|---|---|---|---|---|---|---|---|---|---|---|---|---|---|---|---|---|---|---|---|
| | July | Aug | Sep | Oct | Nov | Dec | Jan | Feb | Mar | Apr | May | June | July | Aug | Sep | Oct | Nov | Dec | Jan | Feb | Mar |
| AY.46.4 | 0 | 0 | 0 | 0 | 0 | 0 | 0 | 0 | 0 | 0 | 0 | 0 | 3 | 10 | 16 | 8 | 1 | 3 | 0 | 0 | 0 |
| AY.47 | 0 | 0 | 0 | 0 | 0 | 0 | 0 | 0 | 0 | 0 | 0 | 0 | 15 | 82 | 96 | 18 | 6 | 2 | 0 | 0 | 0 |
| AY.48 | 0 | 0 | 0 | 0 | 0 | 0 | 0 | 0 | 0 | 0 | 0 | 3 | 6 | 2 | 2 | 0 | 0 | 0 | 0 | 0 | 0 |
| AY.49 | 0 | 0 | 0 | 0 | 0 | 0 | 0 | 0 | 0 | 0 | 0 | 0 | 0 | 0 | 0 | 1 | 0 | 0 | 0 | 0 | 0 |
| AY.5 | 0 | 0 | 0 | 0 | 0 | 0 | 0 | 0 | 0 | 0 | 0 | 0 | 0 | 1 | 0 | 0 | 0 | 0 | 0 | 0 | 0 |
| AY.5.3 | 0 | 0 | 0 | 0 | 0 | 0 | 0 | 0 | 0 | 0 | 0 | 0 | 0 | 0 | 0 | 2 | 0 | 0 | 0 | 0 | 0 |
| AY.52 | 0 | 0 | 0 | 0 | 0 | 0 | 0 | 0 | 0 | 0 | 0 | 2 | 4 | 1 | 0 | 0 | 0 | 0 | 0 | 0 | 0 |
| AY.54 | 0 | 0 | 0 | 0 | 0 | 0 | 0 | 0 | 0 | 0 | 0 | 7 | 35 | 75 | 60 | 3 | 6 | 1 | 1 | 0 | 0 |
| AY.59 | 0 | 0 | 0 | 0 | 0 | 0 | 0 | 0 | 0 | 0 | 0 | 1 | 0 | 2 | 3 | 0 | 0 | 0 | 0 | 0 | 0 |
| AY.62 | 0 | 0 | 0 | 0 | 0 | 0 | 0 | 0 | 0 | 0 | 0 | 0 | 2 | 5 | 1 | 0 | 0 | 0 | 0 | 0 | 0 |
| AY.64 | 0 | 0 | 0 | 0 | 0 | 0 | 0 | 0 | 0 | 0 | 0 | 6 | 1 | 6 | 2 | 0 | 1 | 0 | 0 | 0 | 0 |
| AY.67 | 0 | 0 | 0 | 0 | 0 | 0 | 0 | 0 | 0 | 0 | 0 | 1 | 2 | 3 | 4 | 0 | 1 | 0 | 0 | 0 | 0 |
| AY.7.1 | 0 | 0 | 0 | 0 | 0 | 0 | 0 | 0 | 0 | 0 | 0 | 0 | 0 | 0 | 0 | 1 | 0 | 0 | 0 | 0 | 0 |
| AY.74 | 0 | 0 | 0 | 0 | 0 | 0 | 0 | 0 | 0 | 0 | 0 | 0 | 3 | 1 | 1 | 0 | 0 | 0 | 0 | 0 | 0 |
| AY.75 | 0 | 0 | 0 | 0 | 0 | 0 | 0 | 0 | 0 | 0 | 1 | 0 | 8 | 34 | 42 | 3 | 1 | 3 | 0 | 0 | 0 |
| AY.77 | 0 | 0 | 0 | 0 | 0 | 0 | 0 | 0 | 0 | 0 | 0 | 0 | 0 | 7 | 0 | 0 | 0 | 0 | 0 | 0 | 0 |
| AY.80 | 0 | 0 | 0 | 0 | 0 | 0 | 0 | 0 | 0 | 0 | 0 | 0 | 0 | 1 | 0 | 0 | 0 | 0 | 0 | 0 | 0 |
| AY.81 | 0 | 0 | 0 | 0 | 0 | 0 | 0 | 0 | 0 | 0 | 0 | 3 | 6 | 12 | 11 | 2 | 0 | 0 | 0 | 0 | 0 |
| AY.82 | 0 | 0 | 0 | 0 | 0 | 0 | 0 | 0 | 0 | 0 | 0 | 0 | 0 | 0 | 1 | 0 | 0 | 0 | 0 | 0 | 0 |

Approved for Public Release; Distribution Unlimited. Public Release Case Number 22-2693
© 2023 The MITRE Corporation. All Rights Reserved.

| PANGO Lineage | 2020 | | | | | | 2021 | | | | | | | | | | | | 2022 | | |
|---|---|---|---|---|---|---|---|---|---|---|---|---|---|---|---|---|---|---|---|---|---|
| | July | Aug | Sep | Oct | Nov | Dec | Jan | Feb | Mar | Apr | May | June | July | Aug | Sep | Oct | Nov | Dec | Jan | Feb | Mar |
| AY.83 | 0 | 0 | 0 | 0 | 0 | 0 | 0 | 0 | 0 | 0 | 0 | 0 | 1 | 2 | 1 | 1 | 1 | 0 | 0 | 0 | 0 |
| AY.86 | 0 | 0 | 0 | 0 | 0 | 0 | 0 | 0 | 0 | 0 | 0 | 0 | 0 | 0 | 1 | 1 | 0 | 0 | 0 | 0 | 0 |
| AY.88 | 0 | 0 | 0 | 0 | 0 | 0 | 0 | 0 | 0 | 0 | 0 | 0 | 0 | 1 | 1 | 0 | 0 | 0 | 0 | 0 | 0 |
| AY.9.2 | 0 | 0 | 0 | 0 | 0 | 0 | 0 | 0 | 0 | 0 | 0 | 0 | 0 | 3 | 4 | 3 | 0 | 1 | 1 | 0 | 0 |
| AY.91 | 0 | 0 | 0 | 0 | 0 | 0 | 0 | 0 | 0 | 0 | 0 | 0 | 0 | 1 | 0 | 0 | 0 | 0 | 0 | 0 | 0 |
| AY.92 | 0 | 0 | 0 | 0 | 0 | 0 | 0 | 0 | 0 | 0 | 0 | 0 | 0 | 0 | 0 | 1 | 0 | 0 | 0 | 0 | 0 |
| AY.98.1 | 0 | 0 | 0 | 0 | 0 | 0 | 0 | 0 | 0 | 0 | 0 | 0 | 0 | 1 | 1 | 1 | 5 | 0 | 0 | 0 | 0 |
| AZ.3 | 0 | 0 | 0 | 0 | 0 | 0 | 0 | 0 | 0 | 2 | 2 | 0 | 0 | 0 | 0 | 0 | 0 | 0 | 0 | 0 | 0 |
| B | 0 | 0 | 0 | 0 | 0 | 0 | 0 | 0 | 0 | 0 | 0 | 0 | 0 | 3 | 1 | 0 | 0 | 0 | 0 | 0 | 0 |
| B.1 | 1 | 0 | 0 | 1 | 0 | 9 | 6 | 26 | 20 | 34 | 23 | 15 | 5 | 7 | 2 | 0 | 0 | 2 | 3 | 0 | 0 |
| B.1.1 | 0 | 0 | 0 | 0 | 0 | 1 | 1 | 3 | 2 | 1 | 0 | 1 | 0 | 1 | 0 | 0 | 0 | 0 | 0 | 0 | 0 |
| B.1.1.186 | 0 | 0 | 0 | 0 | 0 | 1 | 0 | 0 | 0 | 0 | 0 | 0 | 0 | 0 | 0 | 0 | 0 | 0 | 0 | 0 | 0 |
| B.1.1.192 | 0 | 0 | 0 | 0 | 0 | 1 | 0 | 0 | 0 | 0 | 0 | 0 | 0 | 0 | 0 | 0 | 0 | 0 | 0 | 0 | 0 |
| B.1.1.222 | 0 | 0 | 0 | 0 | 0 | 3 | 6 | 19 | 1 | 0 | 0 | 0 | 0 | 0 | 0 | 0 | 0 | 0 | 1 | 0 | 0 |
| B.1.1.239 | 0 | 0 | 0 | 0 | 0 | 1 | 0 | 0 | 0 | 0 | 0 | 0 | 0 | 0 | 0 | 0 | 0 | 0 | 0 | 0 | 0 |
| B.1.1.265 | 0 | 0 | 0 | 0 | 0 | 1 | 0 | 0 | 0 | 0 | 0 | 0 | 0 | 0 | 0 | 0 | 0 | 0 | 0 | 0 | 0 |
| B.1.1.318 | 0 | 0 | 0 | 0 | 0 | 0 | 0 | 0 | 0 | 5 | 2 | 2 | 0 | 0 | 0 | 0 | 0 | 0 | 0 | 0 | 0 |
| B.1.1.416 | 0 | 0 | 0 | 0 | 1 | 7 | 3 | 1 | 0 | 0 | 0 | 0 | 0 | 0 | 0 | 0 | 0 | 0 | 0 | 0 | 0 |
| B.1.1.432 | 0 | 0 | 0 | 0 | 0 | 1 | 0 | 2 | 0 | 0 | 0 | 0 | 0 | 0 | 0 | 0 | 0 | 0 | 0 | 0 | 0 |

Approved for Public Release; Distribution Unlimited. Public Release Case Number 22-2693
© 2023 The MITRE Corporation. All Rights Reserved.

| PANGO Lineage | 2020 | | | | | | 2021 | | | | | | | | | | | | 2022 | | |
|---|---|---|---|---|---|---|---|---|---|---|---|---|---|---|---|---|---|---|---|---|---|
| | July | Aug | Sep | Oct | Nov | Dec | Jan | Feb | Mar | Apr | May | June | July | Aug | Sep | Oct | Nov | Dec | Jan | Feb | Mar |
| B.1.1.519 | 0 | 0 | 0 | 0 | 0 | 0 | 6 | 7 | 54 | 48 | 10 | 2 | 0 | 0 | 0 | 0 | 0 | 0 | 0 | 0 | 0 |
| B.1.1.7 | 0 | 0 | 0 | 0 | 0 | 0 | 0 | 32 | 224 | 388 | 304 | 114 | 18 | 2 | 0 | 0 | 0 | 0 | 0 | 0 | 0 |
| B.1.1.8 | 1 | 0 | 0 | 0 | 0 | 0 | 0 | 0 | 0 | 0 | 0 | 0 | 0 | 0 | 0 | 0 | 0 | 0 | 0 | 0 | 0 |
| B.1.110.3 | 0 | 0 | 0 | 0 | 1 | 0 | 0 | 0 | 0 | 0 | 0 | 0 | 0 | 0 | 0 | 0 | 0 | 0 | 0 | 0 | 0 |
| B.1.126 | 0 | 0 | 0 | 0 | 0 | 1 | 0 | 0 | 0 | 0 | 0 | 0 | 0 | 0 | 0 | 0 | 0 | 0 | 0 | 0 | 0 |
| B.1.2 | 0 | 0 | 0 | 0 | 3 | 33 | 71 | 137 | 87 | 28 | 0 | 0 | 0 | 0 | 0 | 0 | 0 | 0 | 0 | 1 | 0 |
| B.1.222 | 0 | 0 | 0 | 0 | 0 | 0 | 1 | 0 | 0 | 0 | 0 | 0 | 0 | 0 | 0 | 0 | 0 | 0 | 0 | 0 | 0 |
| B.1.232 | 0 | 1 | 0 | 0 | 0 | 1 | 1 | 0 | 0 | 0 | 0 | 0 | 0 | 0 | 0 | 0 | 0 | 0 | 0 | 0 | 0 |
| B.1.234 | 0 | 0 | 0 | 0 | 0 | 4 | 1 | 6 | 2 | 0 | 0 | 0 | 0 | 0 | 0 | 0 | 0 | 0 | 0 | 0 | 0 |
| B.1.239 | 0 | 0 | 0 | 0 | 2 | 2 | 0 | 0 | 0 | 0 | 0 | 0 | 0 | 0 | 0 | 0 | 0 | 0 | 0 | 0 | 0 |
| B.1.240 | 0 | 0 | 0 | 0 | 0 | 0 | 1 | 1 | 0 | 0 | 0 | 0 | 0 | 0 | 0 | 0 | 0 | 0 | 0 | 0 | 0 |
| B.1.241 | 0 | 0 | 0 | 0 | 0 | 2 | 0 | 0 | 0 | 0 | 0 | 0 | 0 | 0 | 0 | 0 | 0 | 0 | 0 | 0 | 0 |
| B.1.243 | 0 | 0 | 0 | 0 | 0 | 4 | 2 | 4 | 2 | 0 | 0 | 0 | 0 | 0 | 0 | 0 | 0 | 0 | 0 | 0 | 0 |
| B.1.258 | 0 | 0 | 0 | 0 | 0 | 2 | 0 | 0 | 0 | 0 | 0 | 0 | 0 | 0 | 0 | 0 | 0 | 0 | 0 | 0 | 0 |
| B.1.258.23 | 0 | 0 | 0 | 0 | 0 | 0 | 8 | 0 | 0 | 0 | 0 | 0 | 0 | 0 | 0 | 0 | 0 | 0 | 0 | 0 | 0 |
| B.1.265 | 0 | 0 | 0 | 0 | 0 | 1 | 0 | 0 | 0 | 0 | 0 | 0 | 0 | 0 | 0 | 0 | 0 | 0 | 0 | 0 | 0 |
| B.1.313 | 0 | 0 | 0 | 0 | 0 | 0 | 0 | 0 | 2 | 0 | 1 | 1 | 0 | 0 | 0 | 0 | 0 | 0 | 0 | 0 | 0 |
| B.1.324 | 0 | 0 | 0 | 0 | 0 | 0 | 0 | 0 | 0 | 0 | 1 | 0 | 0 | 0 | 0 | 0 | 0 | 0 | 0 | 0 | 0 |
| B.1.349 | 0 | 0 | 0 | 0 | 0 | 0 | 1 | 0 | 0 | 0 | 0 | 0 | 0 | 0 | 0 | 0 | 0 | 0 | 0 | 0 | 0 |

Approved for Public Release; Distribution Unlimited. Public Release Case Number 22-2693
© 2023 The MITRE Corporation. All Rights Reserved.

| PANGO Lineage | 2020 | | | | | | 2021 | | | | | | | | | | | | 2022 | | |
|---|---|---|---|---|---|---|---|---|---|---|---|---|---|---|---|---|---|---|---|---|---|
| | July | Aug | Sep | Oct | Nov | Dec | Jan | Feb | Mar | Apr | May | June | July | Aug | Sep | Oct | Nov | Dec | Jan | Feb | Mar |
| B.1.351 | 0 | 0 | 0 | 0 | 0 | 0 | 0 | 0 | 0 | 2 | 4 | 3 | 0 | 0 | 0 | 0 | 0 | 0 | 0 | 0 | 0 |
| B.1.36.7 | 0 | 0 | 0 | 0 | 0 | 0 | 0 | 1 | 0 | 0 | 0 | 0 | 0 | 0 | 0 | 0 | 0 | 0 | 0 | 0 | 0 |
| B.1.369 | 0 | 0 | 0 | 0 | 0 | 1 | 1 | 0 | 0 | 0 | 0 | 0 | 0 | 0 | 0 | 0 | 0 | 0 | 0 | 0 | 0 |
| B.1.375 | 0 | 0 | 0 | 0 | 0 | 0 | 1 | 0 | 0 | 0 | 0 | 0 | 0 | 0 | 0 | 0 | 0 | 0 | 0 | 0 | 0 |
| B.1.396 | 0 | 0 | 0 | 0 | 0 | 0 | 0 | 2 | 0 | 0 | 0 | 0 | 0 | 0 | 0 | 0 | 0 | 0 | 0 | 0 | 0 |
| B.1.399 | 0 | 0 | 0 | 0 | 0 | 1 | 0 | 0 | 0 | 0 | 0 | 0 | 0 | 0 | 0 | 0 | 0 | 0 | 0 | 0 | 0 |
| B.1.400 | 0 | 0 | 0 | 0 | 0 | 6 | 9 | 1 | 0 | 0 | 0 | 0 | 0 | 0 | 0 | 0 | 0 | 0 | 0 | 0 | 0 |
| B.1.404 | 0 | 0 | 0 | 0 | 0 | 0 | 1 | 1 | 0 | 0 | 0 | 0 | 0 | 0 | 0 | 0 | 0 | 0 | 0 | 0 | 0 |
| B.1.424 | 0 | 0 | 0 | 0 | 0 | 0 | 0 | 0 | 0 | 0 | 0 | 0 | 0 | 0 | 0 | 0 | 0 | 1 | 0 | 0 | 0 |
| B.1.426 | 0 | 0 | 0 | 0 | 0 | 0 | 1 | 0 | 0 | 0 | 0 | 0 | 0 | 0 | 0 | 0 | 0 | 0 | 0 | 0 | 0 |
| B.1.427 | 0 | 0 | 0 | 0 | 0 | 1 | 11 | 5 | 27 | 18 | 4 | 1 | 0 | 0 | 0 | 0 | 0 | 0 | 0 | 0 | 0 |
| B.1.429 | 0 | 0 | 0 | 0 | 2 | 68 | 85 | 19 | 66 | 36 | 7 | 0 | 1 | 0 | 0 | 0 | 0 | 0 | 0 | 0 | 0 |
| B.1.433 | 0 | 0 | 0 | 0 | 0 | 5 | 0 | 0 | 0 | 0 | 0 | 0 | 0 | 0 | 0 | 0 | 0 | 0 | 0 | 0 | 0 |
| B.1.525 | 0 | 0 | 0 | 0 | 0 | 0 | 0 | 1 | 0 | 0 | 1 | 0 | 0 | 0 | 0 | 0 | 0 | 0 | 0 | 0 | 0 |
| B.1.526 | 0 | 0 | 0 | 0 | 0 | 0 | 0 | 0 | 5 | 4 | 4 | 5 | 1 | 0 | 0 | 0 | 0 | 0 | 0 | 0 | 0 |
| B.1.544 | 0 | 0 | 0 | 0 | 0 | 0 | 0 | 0 | 1 | 0 | 0 | 0 | 0 | 0 | 0 | 0 | 0 | 0 | 0 | 0 | 0 |
| B.1.551 | 0 | 0 | 0 | 0 | 0 | 1 | 1 | 0 | 1 | 0 | 0 | 0 | 0 | 0 | 0 | 0 | 0 | 0 | 0 | 0 | 0 |
| B.1.561 | 0 | 0 | 0 | 0 | 0 | 7 | 1 | 0 | 11 | 4 | 0 | 0 | 0 | 0 | 0 | 0 | 0 | 0 | 0 | 0 | 0 |
| B.1.565 | 0 | 0 | 0 | 0 | 0 | 1 | 0 | 0 | 0 | 0 | 0 | 0 | 0 | 0 | 0 | 0 | 0 | 0 | 0 | 0 | 0 |

Approved for Public Release; Distribution Unlimited. Public Release Case Number 22-2693
© 2023 The MITRE Corporation. All Rights Reserved.

| PANGO Lineage | 2020 | | | | | | 2021 | | | | | | | | | | | | 2022 | | |
|---|---|---|---|---|---|---|---|---|---|---|---|---|---|---|---|---|---|---|---|---|---|
| | July | Aug | Sep | Oct | Nov | Dec | Jan | Feb | Mar | Apr | May | June | July | Aug | Sep | Oct | Nov | Dec | Jan | Feb | Mar |
| B.1.568 | 0 | 0 | 0 | 0 | 0 | 0 | 0 | 1 | 0 | 0 | 0 | 0 | 0 | 0 | 0 | 0 | 0 | 0 | 0 | 0 | 0 |
| B.1.575 | 0 | 0 | 0 | 0 | 0 | 0 | 0 | 1 | 1 | 4 | 9 | 0 | 0 | 0 | 0 | 0 | 0 | 0 | 0 | 0 | 0 |
| B.1.577 | 0 | 0 | 0 | 0 | 0 | 0 | 0 | 1 | 0 | 0 | 0 | 0 | 0 | 0 | 0 | 0 | 0 | 0 | 0 | 0 | 0 |
| B.1.582 | 0 | 0 | 0 | 0 | 0 | 0 | 0 | 1 | 0 | 0 | 0 | 0 | 0 | 0 | 0 | 0 | 0 | 0 | 0 | 0 | 0 |
| B.1.587 | 0 | 0 | 0 | 0 | 0 | 0 | 0 | 2 | 0 | 0 | 0 | 0 | 0 | 0 | 0 | 0 | 0 | 0 | 0 | 0 | 0 |
| B.1.595 | 0 | 0 | 0 | 0 | 0 | 6 | 3 | 1 | 1 | 0 | 1 | 0 | 0 | 0 | 0 | 0 | 0 | 0 | 0 | 0 | 0 |
| B.1.596 | 0 | 0 | 0 | 0 | 0 | 0 | 0 | 1 | 1 | 1 | 0 | 0 | 0 | 0 | 0 | 0 | 0 | 0 | 0 | 0 | 0 |
| B.1.599 | 0 | 0 | 0 | 0 | 0 | 0 | 1 | 0 | 0 | 0 | 0 | 0 | 0 | 0 | 0 | 0 | 0 | 0 | 0 | 0 | 0 |
| B.1.609 | 0 | 0 | 0 | 0 | 0 | 3 | 2 | 3 | 0 | 0 | 0 | 0 | 0 | 0 | 0 | 0 | 0 | 0 | 0 | 0 | 0 |
| B.1.612 | 0 | 0 | 0 | 0 | 0 | 0 | 1 | 0 | 1 | 1 | 0 | 0 | 0 | 0 | 0 | 0 | 0 | 0 | 0 | 0 | 0 |
| B.1.617.1 | 0 | 0 | 0 | 0 | 0 | 0 | 0 | 0 | 0 | 1 | 0 | 0 | 0 | 0 | 0 | 0 | 0 | 0 | 0 | 0 | 0 |
| B.1.617.2 | 0 | 0 | 0 | 0 | 0 | 0 | 0 | 0 | 0 | 0 | 0 | 4 | 9 | 92 | 68 | 35 | 26 | 6 | 1 | 0 | 0 |
| B.1.621 | 0 | 0 | 0 | 0 | 0 | 0 | 0 | 0 | 0 | 4 | 32 | 15 | 9 | 3 | 3 | 0 | 0 | 0 | 0 | 0 | 0 |
| B.1.621.1 | 0 | 0 | 0 | 0 | 0 | 0 | 0 | 0 | 0 | 0 | 0 | 0 | 1 | 0 | 0 | 0 | 0 | 0 | 0 | 0 | 0 |
| B.1.621.2 | 0 | 0 | 0 | 0 | 0 | 0 | 0 | 0 | 0 | 1 | 0 | 0 | 0 | 0 | 0 | 0 | 0 | 0 | 0 | 0 | 0 |
| B.1.627 | 0 | 0 | 0 | 0 | 0 | 0 | 0 | 0 | 1 | 1 | 0 | 0 | 0 | 0 | 0 | 0 | 0 | 0 | 0 | 0 | 0 |
| B.1.631 | 0 | 0 | 0 | 0 | 0 | 0 | 0 | 0 | 0 | 0 | 0 | 0 | 0 | 1 | 0 | 0 | 0 | 0 | 0 | 0 | 0 |
| B.1.632 | 0 | 0 | 0 | 0 | 0 | 0 | 0 | 0 | 0 | 0 | 0 | 0 | 0 | 2 | 0 | 0 | 0 | 0 | 0 | 0 | 0 |
| B.1.637 | 0 | 0 | 0 | 0 | 0 | 0 | 0 | 1 | 1 | 2 | 2 | 1 | 0 | 0 | 0 | 0 | 0 | 0 | 0 | 0 | 0 |

Approved for Public Release; Distribution Unlimited. Public Release Case Number 22-2693
© 2023 The MITRE Corporation. All Rights Reserved.

| PANGO Lineage | 2020 | | | | | | 2021 | | | | | | | | | | | | 2022 | | |
|---|---|---|---|---|---|---|---|---|---|---|---|---|---|---|---|---|---|---|---|---|---|
| | July | Aug | Sep | Oct | Nov | Dec | Jan | Feb | Mar | Apr | May | June | July | Aug | Sep | Oct | Nov | Dec | Jan | Feb | Mar |
| BA.1 | 0 | 0 | 0 | 0 | 0 | 0 | 0 | 0 | 0 | 0 | 0 | 0 | 0 | 0 | 0 | 0 | 0 | 6 | 39 | 1 | 0 |
| BA.1.1 | 0 | 0 | 0 | 0 | 0 | 0 | 0 | 0 | 0 | 0 | 0 | 0 | 0 | 0 | 0 | 0 | 0 | 133 | 1,120 | 204 | 27 |
| BA.1.1.1 | 0 | 0 | 0 | 0 | 0 | 0 | 0 | 0 | 0 | 0 | 0 | 0 | 0 | 0 | 0 | 0 | 0 | 1 | 27 | 2 | 0 |
| BA.1.1.10 | 0 | 0 | 0 | 0 | 0 | 0 | 0 | 0 | 0 | 0 | 0 | 0 | 0 | 0 | 0 | 0 | 0 | 0 | 3 | 0 | 0 |
| BA.1.1.11 | 0 | 0 | 0 | 0 | 0 | 0 | 0 | 0 | 0 | 0 | 0 | 0 | 0 | 0 | 0 | 0 | 0 | 3 | 24 | 4 | 0 |
| BA.1.1.12 | 0 | 0 | 0 | 0 | 0 | 0 | 0 | 0 | 0 | 0 | 0 | 0 | 0 | 0 | 0 | 0 | 0 | 0 | 0 | 1 | 0 |
| BA.1.1.13 | 0 | 0 | 0 | 0 | 0 | 0 | 0 | 0 | 0 | 0 | 0 | 0 | 0 | 0 | 0 | 0 | 0 | 0 | 1 | 0 | 0 |
| BA.1.1.14 | 0 | 0 | 0 | 0 | 0 | 0 | 0 | 0 | 0 | 0 | 0 | 0 | 0 | 0 | 0 | 0 | 0 | 6 | 12 | 4 | 0 |
| BA.1.1.15 | 0 | 0 | 0 | 0 | 0 | 0 | 0 | 0 | 0 | 0 | 0 | 0 | 0 | 0 | 0 | 0 | 0 | 1 | 9 | 0 | 0 |
| BA.1.1.16 | 0 | 0 | 0 | 0 | 0 | 0 | 0 | 0 | 0 | 0 | 0 | 0 | 0 | 0 | 0 | 0 | 0 | 4 | 10 | 0 | 1 |
| BA.1.1.2 | 0 | 0 | 0 | 0 | 0 | 0 | 0 | 0 | 0 | 0 | 0 | 0 | 0 | 0 | 0 | 0 | 0 | 0 | 17 | 1 | 0 |
| BA.1.1.4 | 0 | 0 | 0 | 0 | 0 | 0 | 0 | 0 | 0 | 0 | 0 | 0 | 0 | 0 | 0 | 0 | 0 | 0 | 1 | 0 | 0 |
| BA.1.1.5 | 0 | 0 | 0 | 0 | 0 | 0 | 0 | 0 | 0 | 0 | 0 | 0 | 0 | 0 | 0 | 0 | 0 | 0 | 1 | 0 | 0 |
| BA.1.1.8 | 0 | 0 | 0 | 0 | 0 | 0 | 0 | 0 | 0 | 0 | 0 | 0 | 0 | 0 | 0 | 0 | 0 | 1 | 2 | 0 | 0 |
| BA.1.13 | 0 | 0 | 0 | 0 | 0 | 0 | 0 | 0 | 0 | 0 | 0 | 0 | 0 | 0 | 0 | 0 | 0 | 0 | 7 | 1 | 0 |
| BA.1.14 | 0 | 0 | 0 | 0 | 0 | 0 | 0 | 0 | 0 | 0 | 0 | 0 | 0 | 0 | 0 | 0 | 0 | 35 | 129 | 18 | 1 |
| BA.1.15 | 0 | 0 | 0 | 0 | 0 | 0 | 0 | 0 | 0 | 0 | 0 | 0 | 0 | 0 | 0 | 0 | 0 | 405 | 463 | 32 | 3 |
| BA.1.15.1 | 0 | 0 | 0 | 0 | 0 | 0 | 0 | 0 | 0 | 0 | 0 | 0 | 0 | 0 | 0 | 0 | 0 | 1 | 35 | 0 | 0 |
| BA.1.15.2 | 0 | 0 | 0 | 0 | 0 | 0 | 0 | 0 | 0 | 0 | 0 | 0 | 0 | 0 | 0 | 0 | 0 | 4 | 2 | 0 | 0 |

Approved for Public Release; Distribution Unlimited. Public Release Case Number 22-2693
© 2023 The MITRE Corporation. All Rights Reserved.

| PANGO Lineage | 2020 | | | | | | 2021 | | | | | | | | | | | | 2022 | | |
|---|---|---|---|---|---|---|---|---|---|---|---|---|---|---|---|---|---|---|---|---|---|
| | July | Aug | Sep | Oct | Nov | Dec | Jan | Feb | Mar | Apr | May | June | July | Aug | Sep | Oct | Nov | Dec | Jan | Feb | Mar |
| BA.1.16 | 0 | 0 | 0 | 0 | 0 | 0 | 0 | 0 | 0 | 0 | 0 | 0 | 0 | 0 | 0 | 0 | 0 | 7 | 35 | 0 | 0 |
| BA.1.17 | 0 | 0 | 0 | 0 | 0 | 0 | 0 | 0 | 0 | 0 | 0 | 0 | 0 | 0 | 0 | 0 | 0 | 1 | 12 | 1 | 0 |
| BA.1.17.2 | 0 | 0 | 0 | 0 | 0 | 0 | 0 | 0 | 0 | 0 | 0 | 0 | 0 | 0 | 0 | 0 | 0 | 2 | 18 | 0 | 0 |
| BA.1.18 | 0 | 0 | 0 | 0 | 0 | 0 | 0 | 0 | 0 | 0 | 0 | 0 | 0 | 0 | 0 | 0 | 0 | 7 | 18 | 1 | 0 |
| BA.1.19 | 0 | 0 | 0 | 0 | 0 | 0 | 0 | 0 | 0 | 0 | 0 | 0 | 0 | 0 | 0 | 0 | 0 | 3 | 4 | 2 | 0 |
| BA.1.20 | 0 | 0 | 0 | 0 | 0 | 0 | 0 | 0 | 0 | 0 | 0 | 0 | 0 | 0 | 0 | 0 | 0 | 68 | 112 | 70 | 1 |
| BA.1.21 | 0 | 0 | 0 | 0 | 0 | 0 | 0 | 0 | 0 | 0 | 0 | 0 | 0 | 0 | 0 | 0 | 0 | 2 | 1 | 0 | 0 |
| BA.1.5 | 0 | 0 | 0 | 0 | 0 | 0 | 0 | 0 | 0 | 0 | 0 | 0 | 0 | 0 | 0 | 0 | 0 | 0 | 2 | 0 | 0 |
| BA.1.6 | 0 | 0 | 0 | 0 | 0 | 0 | 0 | 0 | 0 | 0 | 0 | 0 | 0 | 0 | 0 | 0 | 0 | 0 | 0 | 1 | 0 |
| BA.1.9 | 0 | 0 | 0 | 0 | 0 | 0 | 0 | 0 | 0 | 0 | 0 | 0 | 0 | 0 | 0 | 0 | 0 | 1 | 4 | 0 | 0 |
| BA.2 | 0 | 0 | 0 | 0 | 0 | 0 | 0 | 0 | 0 | 0 | 0 | 0 | 0 | 0 | 0 | 0 | 0 | 0 | 1 | 0 | 1 |
| BA.2.3 | 0 | 0 | 0 | 0 | 0 | 0 | 0 | 0 | 0 | 0 | 0 | 0 | 0 | 0 | 0 | 0 | 0 | 0 | 1 | 2 | 0 |
| BA.2.5 | 0 | 0 | 0 | 0 | 0 | 0 | 0 | 0 | 0 | 0 | 0 | 0 | 0 | 0 | 0 | 0 | 0 | 1 | 0 | 3 | 0 |
| BA.2.9 | 0 | 0 | 0 | 0 | 0 | 0 | 0 | 0 | 0 | 0 | 0 | 0 | 0 | 0 | 0 | 0 | 0 | 0 | 0 | 2 | 1 |
| C.36 | 0 | 0 | 0 | 0 | 0 | 0 | 0 | 0 | 1 | 0 | 0 | 0 | 0 | 0 | 0 | 0 | 0 | 0 | 0 | 0 | 0 |
| C.37 | 0 | 0 | 0 | 0 | 0 | 0 | 0 | 0 | 0 | 3 | 1 | 0 | 0 | 0 | 0 | 0 | 0 | 0 | 0 | 0 | 0 |
| P.1 | 0 | 0 | 0 | 0 | 0 | 0 | 0 | 0 | 3 | 5 | 19 | 30 | 12 | 2 | 0 | 0 | 0 | 0 | 0 | 0 | 0 |
| P.1.1 | 0 | 0 | 0 | 0 | 0 | 0 | 0 | 0 | 1 | 0 | 1 | 1 | 0 | 0 | 0 | 0 | 0 | 0 | 0 | 0 | 0 |
| P.1.10 | 0 | 0 | 0 | 0 | 0 | 0 | 0 | 0 | 0 | 0 | 1 | 2 | 0 | 0 | 0 | 0 | 0 | 0 | 0 | 0 | 0 |

Approved for Public Release; Distribution Unlimited. Public Release Case Number 22-2693
© 2023 The MITRE Corporation. All Rights Reserved.

| PANGO Lineage | 2020 | | | | | | 2021 | | | | | | | | | | | | 2022 | | |
|---|---|---|---|---|---|---|---|---|---|---|---|---|---|---|---|---|---|---|---|---|---|
| | July | Aug | Sep | Oct | Nov | Dec | Jan | Feb | Mar | Apr | May | June | July | Aug | Sep | Oct | Nov | Dec | Jan | Feb | Mar |
| P.1.14 | 0 | 0 | 0 | 0 | 0 | 0 | 0 | 0 | 0 | 0 | 0 | 2 | 0 | 0 | 0 | 0 | 0 | 0 | 0 | 0 | 0 |
| P.1.15 | 0 | 0 | 0 | 0 | 0 | 0 | 0 | 0 | 0 | 0 | 0 | 1 | 0 | 0 | 0 | 0 | 0 | 0 | 0 | 0 | 0 |
| P.1.17 | 0 | 0 | 0 | 0 | 0 | 0 | 0 | 0 | 0 | 0 | 0 | 0 | 1 | 1 | 0 | 0 | 0 | 0 | 0 | 0 | 0 |
| P.2 | 0 | 0 | 0 | 0 | 0 | 0 | 0 | 4 | 4 | 1 | 0 | 0 | 0 | 0 | 0 | 0 | 0 | 0 | 0 | 0 | 0 |
| Q.3 | 0 | 0 | 0 | 0 | 0 | 0 | 0 | 0 | 3 | 12 | 5 | 3 | 1 | 0 | 0 | 0 | 0 | 0 | 0 | 0 | 0 |
| Q.8 | 0 | 0 | 0 | 0 | 0 | 0 | 0 | 0 | 0 | 1 | 0 | 0 | 0 | 0 | 0 | 0 | 0 | 0 | 0 | 0 | 0 |
| R.1 | 0 | 0 | 0 | 0 | 0 | 0 | 0 | 2 | 0 | 0 | 0 | 0 | 0 | 0 | 0 | 0 | 0 | 0 | 0 | 0 | 0 |
| XB | 0 | 0 | 0 | 0 | 0 | 0 | 0 | 0 | 0 | 1 | 1 | 0 | 0 | 0 | 0 | 0 | 0 | 0 | 0 | 0 | 0 |
| Total | 2 | 1 | 0 | 1 | 9 | 175 | 226 | 286 | 523 | 611 | 470 | 579 | 765 | 2,814 | 2,731 | 814 | 985 | 1,200 | 2,218 | 351 | 35 |

Approved for Public Release; Distribution Unlimited. Public Release Case Number 22-2693
© 2023 The MITRE Corporation. All Rights Reserved.

### NOTICE

This (software/technical data) was produced for the U. S. Government under Contract Number 75FCMC18D0047, and is subject to Federal Acquisition Regulation Clause 52.227-14, Rights in Data-General.

No other use other than that granted to the U. S. Government, or to those acting on behalf of the U. S. Government under that Clause is authorized without the express written permission of The MITRE Corporation.

For further information, please contact The MITRE Corporation, Contracts Management Office, 7515 Colshire Drive, McLean, VA 22102-7539, (703) 983-6000.

**© 2023 The MITRE Corporation.**

Approved for Public Release; Distribution Unlimited. Public Release Case Number 22-2693
© 2023 The MITRE Corporation. All Rights Reserved.
